# Management of Central Post-Stroke Pain: Systematic Review and Meta-Analysis

**DOI:** 10.1101/2024.01.15.24301311

**Authors:** Arnas Tamasauskas, Andrew Marshall, Barbara Silva-Passadouro, Nichollas Fallon, Bernhard Frank, Svajune Laurinaviciute, Simon Keller

## Abstract

Central post stroke pain (CPSP) is a neuropathic pain condition prevalent in 8% to 35% of stroke patients. This systematic review and meta-analysis aimed to provide insight in the effectiveness of available pharmacological, physical, psychological, and neuromodulation intervention in reducing pain in CPSP patients. Secondary outcomes included mood, sleep, global impression of change, and physical responses. Data extraction included participant demographics, stroke aetiology, pain characteristics, pain reduction scores, and secondary outcome metrics. Forty two original studies were included with a total of 1451 participants. Twelve studies met requirements for a random-effects meta-analysis which found: neuromodulation to be the most effective with a moderate effect on pain scores (SMD = -0.60, 95% confidence interval [-0.97, -0.23]), followed by physical interventions with moderate effect (SMD = -0.55, [-1.28, 0.18), and pharmacological interventions with a small effect on pain (SMD = -0.36, [-0.68, -0.03]). Fourteen studies were included in proportional meta-analysis with pharmacological studies having a moderate effect (58.34% mean reduction, [36.50, 80.18]), and neuromodulation studies a small effect (31.70% mean reduction, [21.44, 41.96]). Sixteen studies were included in the narrative review. While the overall medium risk of bias limits generalisation of findings, fluvoxamine and repetitive transcranial magnetic stimulation was found to have consistently good pain alleviation and relatively low risk of side effects. Anticonvulsants were found to have a significant effect on pain reduction, but were found to have the most side effects. Virtual reality and acupuncture show promising results, but lack rigorous methodological investigation to understand their full effect.

## 1 Introduction

Central Post Stroke Pain (CPSP) is a neuropathic pain condition evident in 8% to 35% of stroke patients[48,57]. CPSP is characterised by spontaneous pain and abnormal somatosensory reaction to sensations, such as: touch, vibration, heat and cold. Pain severity reported by patients ranges from moderate to severe, but on average CPSP patients reported their pain intensity as 6 on an 11 point Visual Analogue Scale (VAS)[28,42]. The onset and qualities of pain can differ between CPSP patients. Spontaneous pain is most common and found in 85% of patients[44]. This type of pain can be either continuous or Paroxysmal, and is described as burning, aching, pricking, or shooting. CPSP patients may also experience evoked pain in conjunction with spontaneous pain or in absence of it. Evoked pain can present as either hyperalgesia to nociceptive stimuli, or allodynia to non-nociceptive thermal or touch stimuli.

Conversely, CPSP may result in negative sensory symptoms with a reduced sensation to tactile and thermal stimuli. Up to 66% of CPSP patients experience impaired perception of pinprick, touch, and thermal stimuli[48]. A smaller portion of CPSP patients also experience impairment of vibration sensation perception. As patients exhibit varied combinations of positive and negative sensory signs, CPSP can be difficult to diagnose.

It has been suggested that a CPSP diagnosis requires the patient to have developed pain after stroke, have experienced a central nervous system lesion as evidenced by brain imaging, and exclusion of other neuropathic causes of pain and non-neuropathic post stroke pain[87]. However, the diagnostic criteria vary in practice with some clinicians not requiring exclusion of other pain conditions, and other clinicians requiring additional criteria, such as including abnormal sensory testing to temperature[47].

The onset of CPSP is also variable with symptoms appearing imminently after stroke or a few years after stroke[57]. Coupled with variable diagnostic criteria, inconstant onset times result in treatment delays[28]. Early CPSP research placed great emphasis on Thalamic lesions as a cause for CPSP and referred to CPSP as ‘Thalamic Syndrome’. However, this was found to be an oversimplification as not all patients with Thalamic lesions developed CPSP, and, conversely, not all patients with CPSP had Thalamic lesions[28]. The presence of a lesion in the spinothalamic pathway does seem to increase the likelihood of developing CPSP but the exact pathomechanisms of this condition are still being investigated[47].

CPSP patients are also at a higher risk of developing physical and psychological comorbidities. One prevalence study reported the presence of non-CPSP pain in over 57% of CPSP patients[45]. This included headaches, shoulder pain, and pain due to muscle spasms. Among psychological comorbidities, CPSP patients experience higher levels of depression, insomnia, drug dependency, and suicide risk[28,48]. These comorbidities were more prevalent in patients with CPSP than in stroke patients without CPSP or stroke patients with dysaesthesia[4]. Albeit, the evidence for this is from relatively small prevalence studies, and the association of CPSP and comorbidities requires further investigation.

Currently, CPSP is regarded as a long-term condition with some speculation that it is life-long[44]. There are no successful disease modifying treatments, but a number of approaches have been explored to reduce CPSP symptoms. From a population-based study, antidepressants were found to be the most common treatment for CPSP as tricyclic antidepressants and serotonin reuptake inhibitors were found, to an extent, to alleviate pain and associated psychological comorbidities[24]. Antidepressants followed by paracetamol and opioids were the commonly used pain relief. Antiepileptics were the third most popular pharmacological treatment with about 25% of participants taking some type of antiepileptic. However, the effectiveness of some pharmacological treatments has been debated. In a systematic review of pharmacological CPSP interventions, Amitriptyline and Lamotrigine were found to be the only oral drugs to provide effective CPSP symptom alleviation but they may also result in unpleasant side effects[24].

Alongside pharmacological interventions, patients with CPSP have also been treated with neuromodulation therapy, physical therapy, and psychological therapy[66,77] with varied effectiveness. The most common neuromodulation techniques for CPSP, invasive Deep Brain Stimulation (DBS) and non-invasive repetitive Transcranial Magnetic Stimulation (rTMS), have mixed effectiveness rates[77,33]. Physical therapy is less common but mirror therapy and acupuncture have been trialled for CPSP treatment[15,18,89]. Finally, cognitive behaviour therapy (CBT) has been prescribed in some instances to help with psychological comorbidity of CPSP[10]. While some clinicians favour one type of therapy over another, treatments are often prescribed together in varying combinations. Literature reviews and guidelines of CPSP treatments are either small in scale, outdated, or focus on a single treatment modality[14,24,77]. Systematic reviews and meta-analysis are also few and limited. Therefore, this systematic review and meta-analysis will build on the existing literature to provide a comprehensive review of CPSP treatment and research studies. The specific aims of this paper will be to (1) collate and describe available neuromodulating, pharmacological, physical and psychological treatments, (2) determine effectiveness of each treatment type, (3) and compare effectiveness of therapies within each treatment type.

## 2 Methods

### 2.1 Protocol Registration

This review paper followed Reporting Items for Systematic Reviews and Meta-Analyses (PRISMA)^63^. The protocol of this review was registered with PROSPERO prior to literature review (CRD42022371835)[83].

### 2.2 Data sources and Searches

A systematic search of electronic databases was undertaken for interventional studies without a set time period with the primary or secondary outcome being reduction in pain for CPSP patients. A full breakdown of databases and journals for hand searching is provided in *Table 1*.

**Table 1:** Searched Databases and Journals.

| Databases | Journals |
| --- | --- |
| CINAHL | Disability and Rehabilitation |
| Cochrane | Patient Education and Counselling |
| Frontiers | International Journal of Stroke |
| Google Scholar | Journal of Neurology |
| PsycINFO | Journal of Pain |
| PubMed | Journal of Pain and Symptom Management |
| Sage | Journal of Stroke and Cerebrovascular Diseases<br>Neurosurgery and Psychiatry |
| Springer | Journal of Stroke |
| Taylor and Francis | Journal of Pain Research |
| Web of Science |  |

The search terms used in the study were formulated using Patient, Intervention, Comparison, and Outcome (PICO) framework[32]. Different wordings or treatment methods within a treatment type were combined using the “OR” operator. The sample and phenomenon of interest key words related to different names of CPSP (“Central Post*Stroke Pain”, “Chronic Post*Stroke Pain”, “Thalamic Pain”, “Dejerine*Roussy syndrome”) and were combined using “AND” operator with design and evaluation terms relating to each treatment type: brain modulating (“Brain modulation”, “Neuromodulation”, “Repetitive Transcranial Magnetic Stimulation”, “TMS”, “rTMS”, “Deep Brain Stimulation”, “DBS”, “Vestibular Stimulation”) pharmacological (“pharmacological”, “Antidepressant”, “Anticonvulsant”, “Opioid”), physical (“Physical”, “Virtual Reality”, “VR”, “Physiotherapy”, “Acupuncture”) and psychological treatments (“Psychological”, “Mindfulness”, “Cognitive Behaviour Therapy”, “CBT”). References of all included papers were searched for any additional studies.

### 2.3 Study Selection

All identified studies were collated using Rayyan[70] systematic review management tool. Titles and abstracts for all studies (n = 5565) were blindly and independently screened by two reviewers (AT and BSP). Any conflict cases were consulted and resolved with a third reviewer (AM). Interrater reliability was calculated using Cohen’s Kappa score.

The inclusion criteria for type of studies to be included were: (1) randomised and non-randomised trials, (2) longitudinal and cross-sectional studies, (3) within and between group controls. The exclusion criteria for the type of studies were: (1) qualitative studies, (2) case studies, (3) reviews, (4) book chapters or articles, (5) animal studies, (6) dissertations, (7) abstract only articles, (8) papers in non-English language, and (9) grey literature. For grey literature pieces which could fit the inclusion criteria, authors were contacted for details of publications. Studies which included multiple chronic pain conditions were included, granted that data relating specifically to CPSP participants was extractable. Studies were excluded if dosage was not disclosed, virtual reality (VR) environments were not defined, or neuromodulation parameters not stated.

Studies were first title screened and then abstract screened. Studies that did not meet the inclusion and exclusion criteria were removed, and the remaining studies were subjected to a full text review. Studies which used “thalamic pain” or “post stroke pain” were reviewed with the third reviewer (AM) to confirm whether the condition was CPSP or a different neuropathic pain condition.

### 2.4 Data Extraction

Once final papers for inclusion were agreed on, a data extraction tool was created by reviewers using Microsoft (US) Excel[61]. Data extraction included mean VAS score change, percentage VAS score change, standard deviations (SD), confidence intervals, and demographic information. Where studies reported multiple outcome timepoints, VAS score change for the last time point was used. Details of all variables extracted from articles are presented in *Table 2*.

**Table 2:**
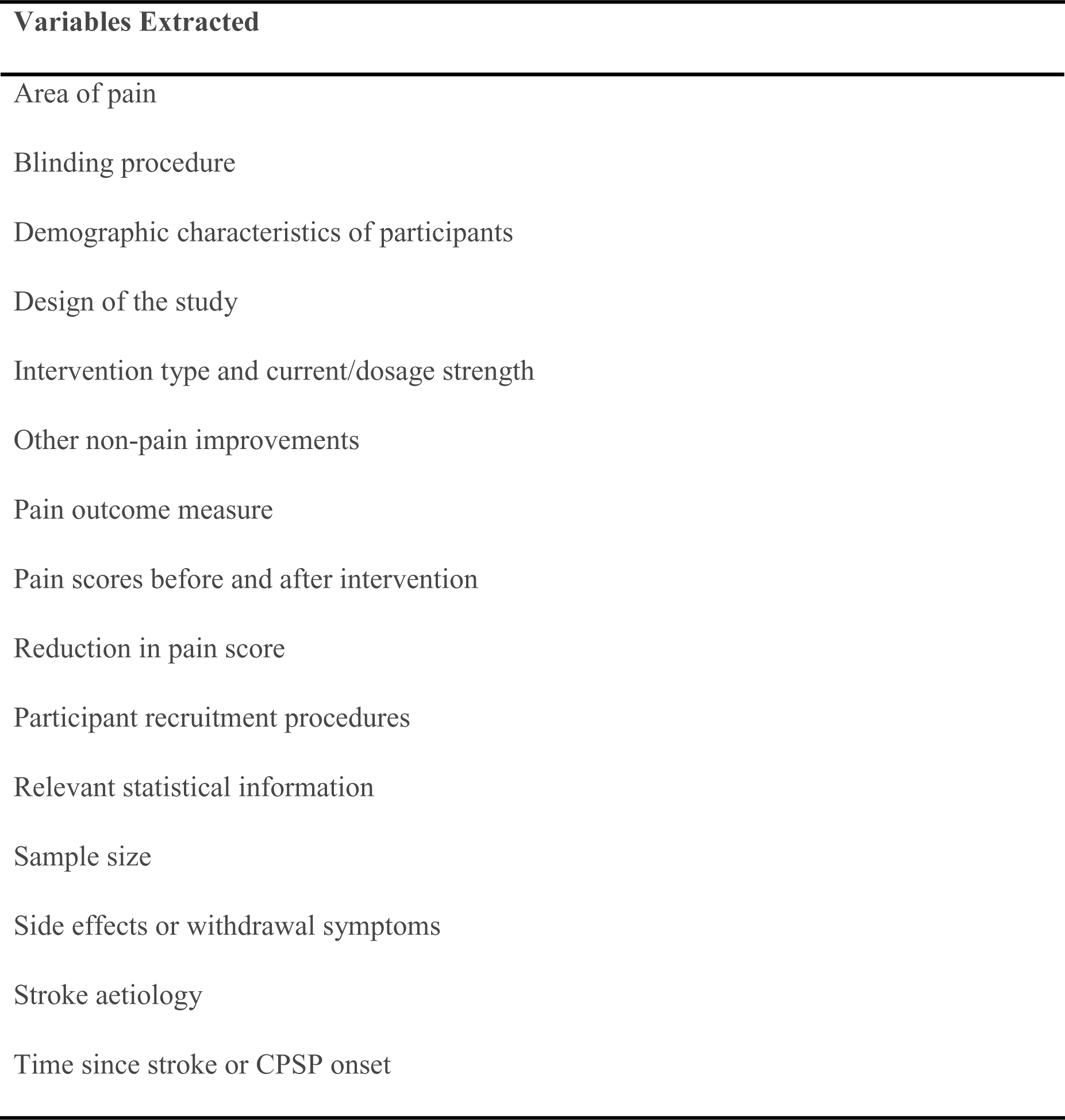
Information variables extracted from included studies.

For studies which did not report standard deviation, mean VAS change or mean percentage change, authors were contacted for information. Out of 15 studies, 6 had listed expired contact information and 7 contacted authors did not respond. Where only an SD score was missing, it was imputed using other available metrics and statistical calculations outlined in Cochrane Handbook for Systematic Reviews of Interventions[29]. Where a mean was not reported but all VAS score or percentage means were provided in the paper, the mean and SD were calculated by reviewers (AT and SL) following a systematic review methodology paper[92]. Studies which had multiple treatment methods or groups with sufficiently different approaches, such as rTMS studies with different target locations or different medications covered in one study, were divided for meta-analysis using calculations from Cochrane Handbook[92]. The approach of splitting the sample size of the main group into multiple groups with smaller sample sizes was adopted for this. Other approaches were considered, such as combining treatment groups into one, but in the interest of covering as many treatment methodologies as possible, these approaches were not adopted.

### 2.5 Quality and Risk Assessment

All studies were quality and risk of bias assessed by two reviewers blindly and independently (AT and SL). Cross sectional studies which were not randomised trials were assessed using the Newcastle-Ottawa Scale (NOS)[93]. Randomised control trials and crossover trials were assessed using a revised Cochrane risk-of-bias tool (RoB 2)[30] and a RoB 2 tool for crossover trials respectively^31^. The respective tools ensured that the quality and risk assessment is appropriate for the study methods.

All studies were additionally scored using the GRADE framework[32] by two independent reviewers (AT and SL). Randomised controlled trials started with high confidence while all other studies started with low confidence. Studies were considered against PICO criteria on imprecision, indirectness, inconsistency, publication bias, and risk of bias (*Table 3*). For each criteria, the study could be downgraded up to 2 times. There were also three positive criteria which could increase the rating of the study: (1) whether different dosage was tested, (2) whether an effect size was calculated and reported, and (3) whether the study considered confounders.

**Table 3:**
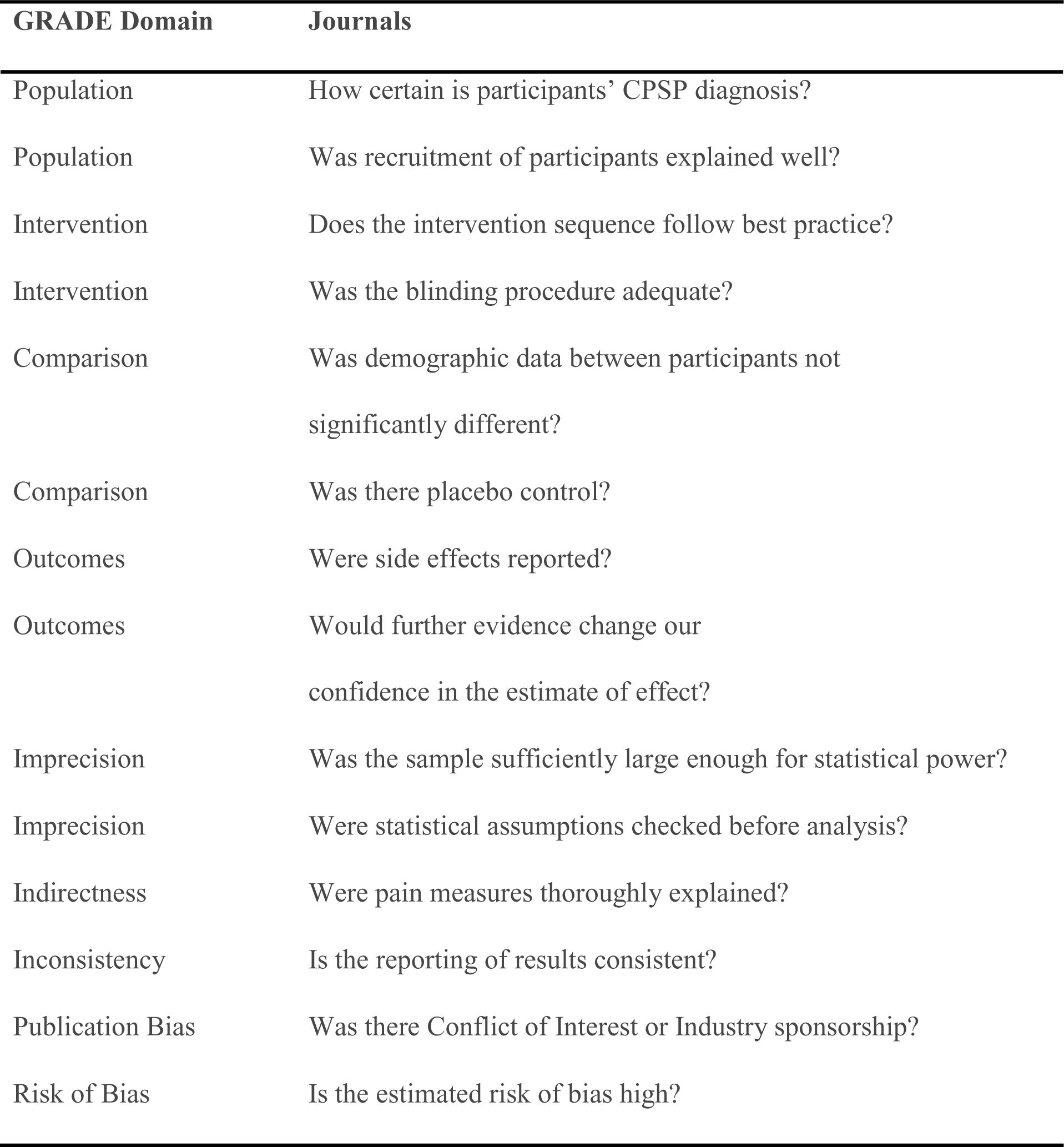
GRADE Framework Negative Criteria.

| GRADE Domain | Journals |
| --- | --- |
| Population | How certain is participants' CPSP diagnosis? |
| Population | Was recruitment of participants explained well? |
| Intervention | Does the intervention sequence follow best practice? |
| Intervention | Was the blinding procedure adequate? |
| Comparison | Was demographic data between participants not significantly different? |
| Comparison | Was there placebo control? |
| Outcomes | Were side effects reported? |
| Outcomes | Would further evidence change our confidence in the estimate of effect? |
| Imprecision | Was the sample sufficiently large enough for statistical power? |
| Imprecision | Were statistical assumptions checked before analysis? |
| Indirectness | Were pain measures thoroughly explained? |
| Inconsistency | Is the reporting of results consistent? |
| Publication Bias | Was there Conflict of Interest or Industry sponsorship? |
| Risk of Bias | Is the estimated risk of bias high? |

### 2.6 Meta Analysis

Data extraction revealed that only 12 studies reported VAS mean score difference and SD metrics for both intervention and comparator groups. This was a limitation that excluded 10 studies from a random-effects model meta-analysis. As this is a field where there is a lack of rigorous comparative studies and summative CPSP treatment reviews, a proportional meta-analysis was conducted on studies that could not be included in the random-effects model. This decision was based on a proportional meta-analysis guide[8] and existing proportional model meta-analysis studies[9,22,25].

Meta-analysis was performed on each treatment group: physical, pharmacological, and neuromodulating. All random-effects meta-analyses were run using R Project using meta library[75,78], while the proportional meta-analyses were run using OpenMeta software[72]. As there was variability in treatment types within each group, select subgroups of specific treatments were analysed. Only treatments with two or more studies were considered for a subgroup meta-analysis. Funnel plot and Egger’s test were performed on all meta-analyses to identify publication bias.

Effect sizes for all meta-analyses were interpreted using SMD and 95% Confidence Intervals. Findings from studies high in risk of bias and or having scored low on quality assessment were considered but these findings should be interpreted cautiously.

## 3 Results

### 3.1 Results of the Search

The systematic database and journal search found 14233 articles. Once duplicates were deleted, 5565 unique studies were identified. After a review of the study titles, 4483 journal articles had their abstracts screened, and 509 of those were selected for full text review. There were 22 studies which resulted in conflicting reviewer opinions. The main conflict was ambiguity around CPSP patient diagnosis. After deliberation and consultation with a third reviewer (AM), it was decided to include 19 of those studies. The search was rerun after 6 months which resulted in one additional study being identified for inclusion. There were 7 studies that did not have CPSP-specific data but met all other inclusion criteria. Authors of these studies were contacted for data but only 1 provided it, while other studies were excluded[4,5,34,42,49,51,65]. Finally, 42 journal articles were identified which met all inclusion criteria. PRISMA flowchart of the review process is detailed in *Figure 1*. The interrater reliability for both reviewers was found to be Kappa = 0.645 (p = 0.04).

**Figure 1:**
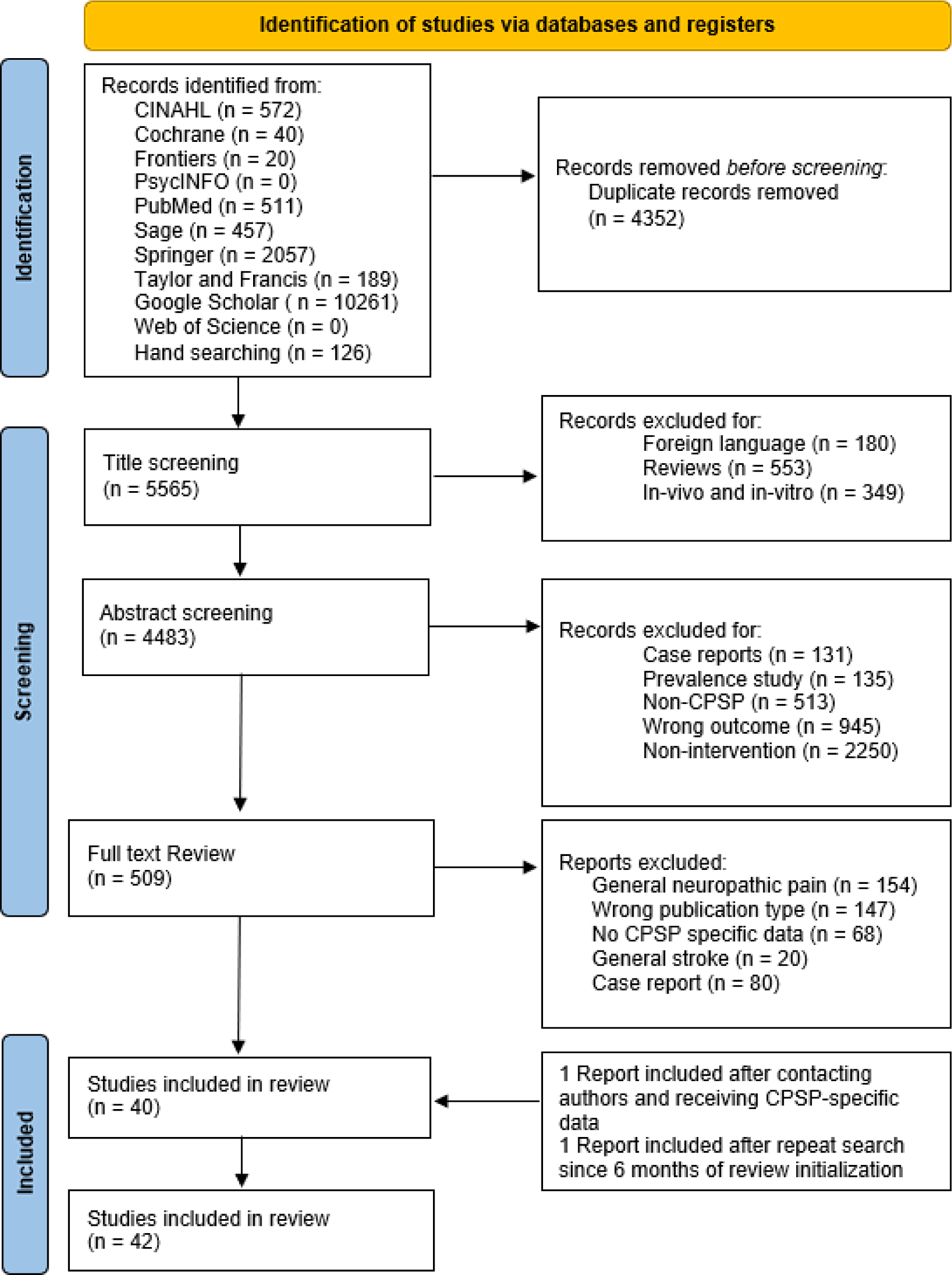
PRISMA flowchart of literature review.

Out of the 42 studies, 12 were randomised control trials, 8 were randomised crossover trials, 5 were retrospective, and 18 were cross sectional interventions. There were 14 pharmacological intervention studies, 5 physical intervention studies, 25 neuromodulation studies, but no psychological interventions were found. Characteristics of studies which were included in either random-effects or proportional meta-analyses are reported in *Table 4*, full study characteristics are provided in *Supplementary Table 1*

**Table 4.**
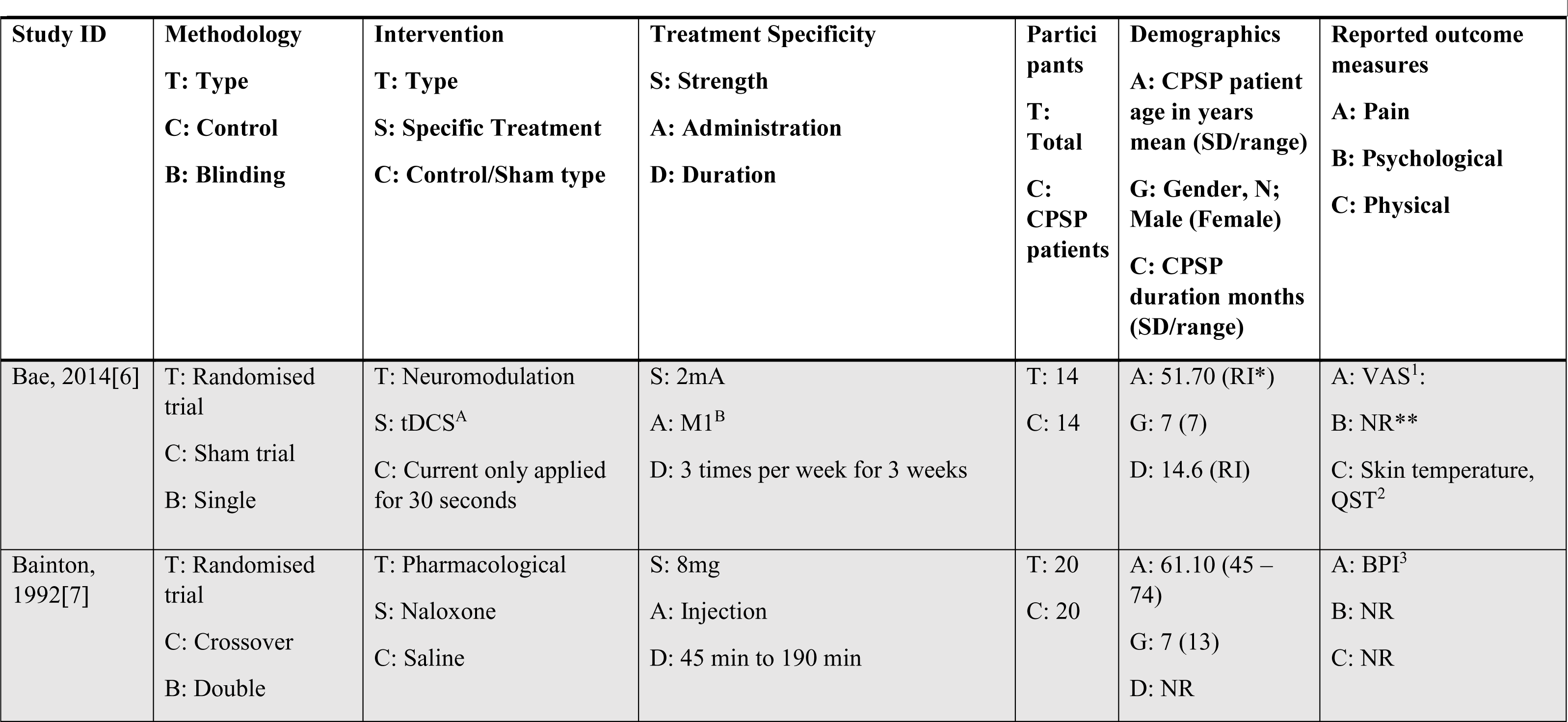
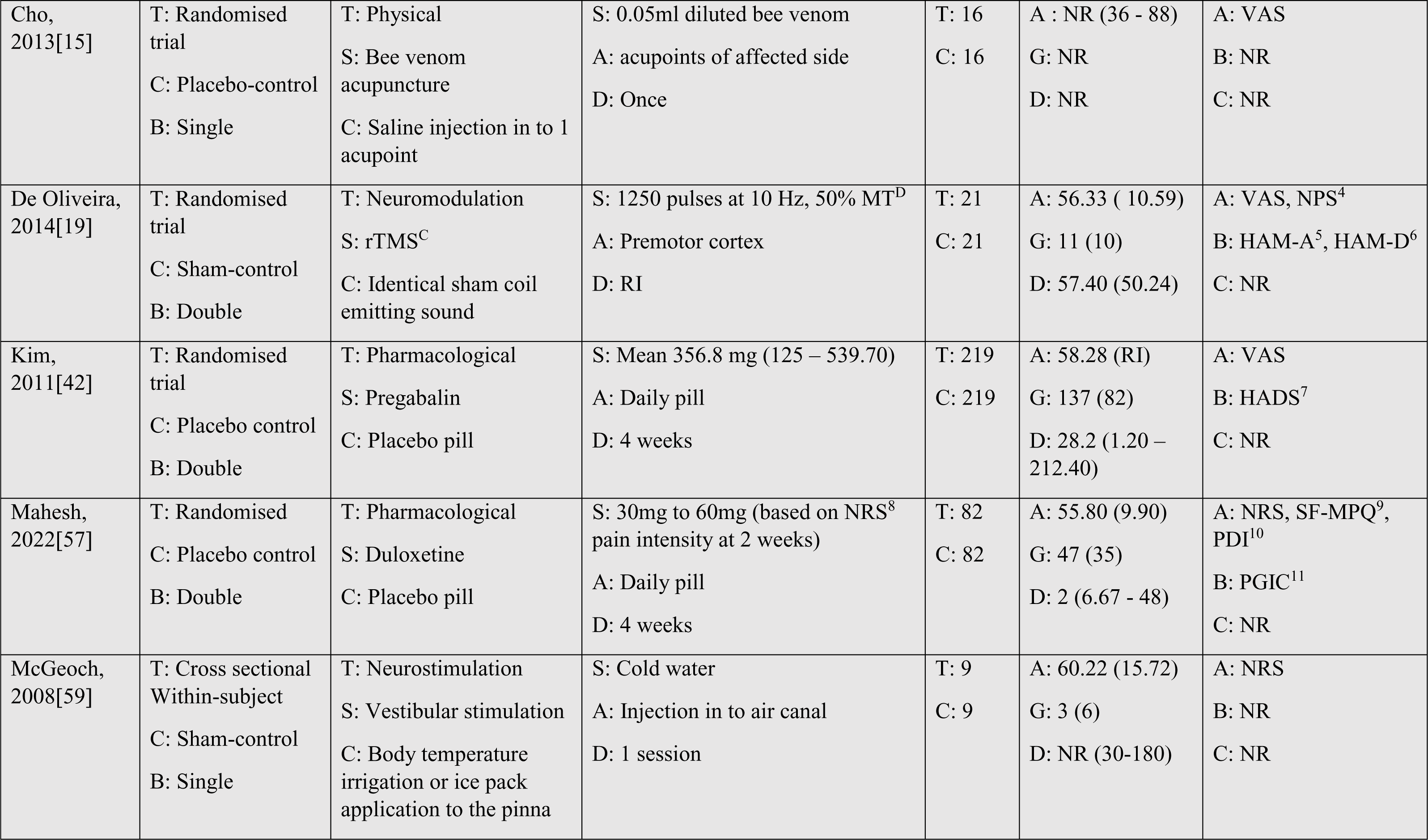
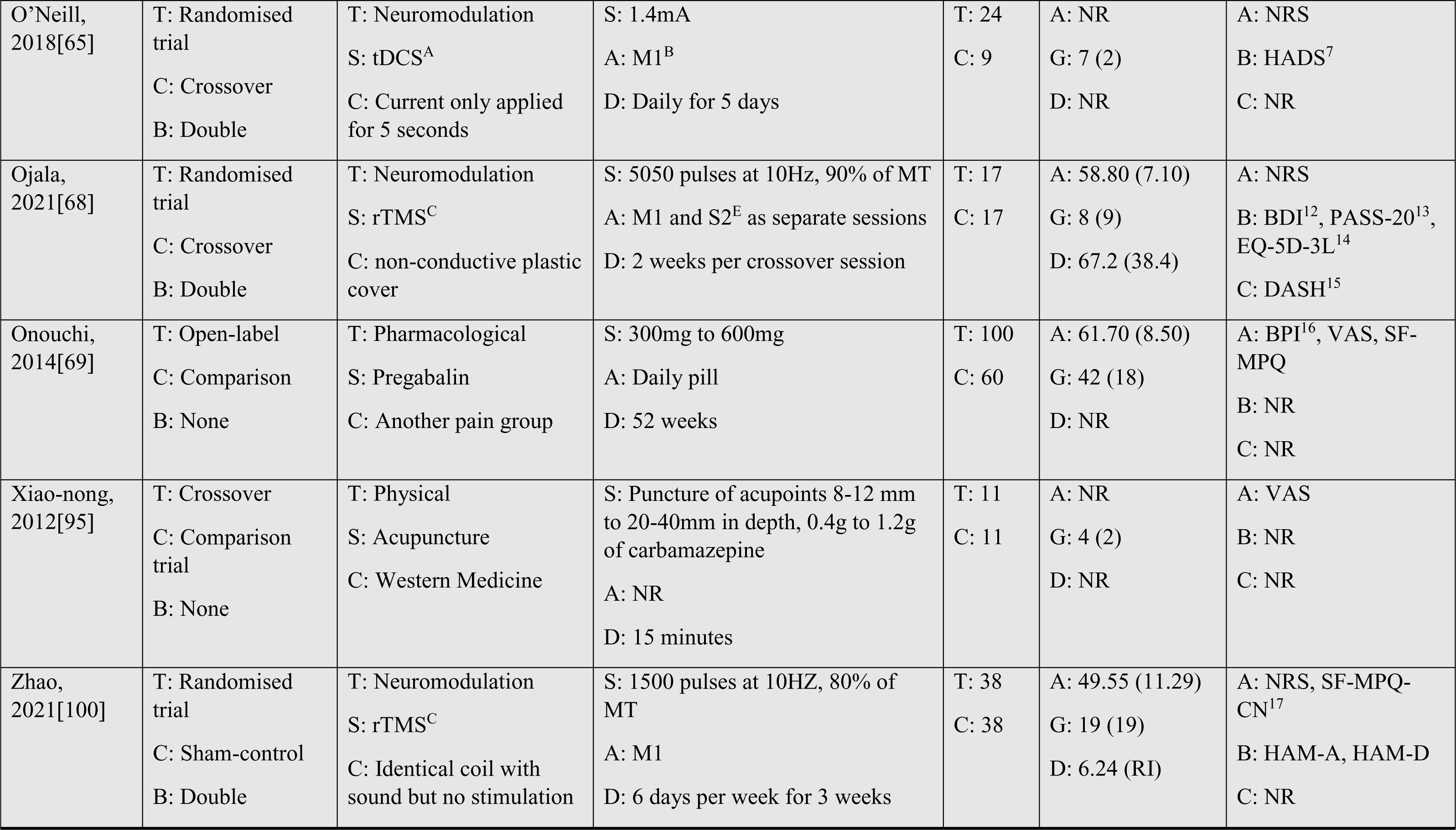
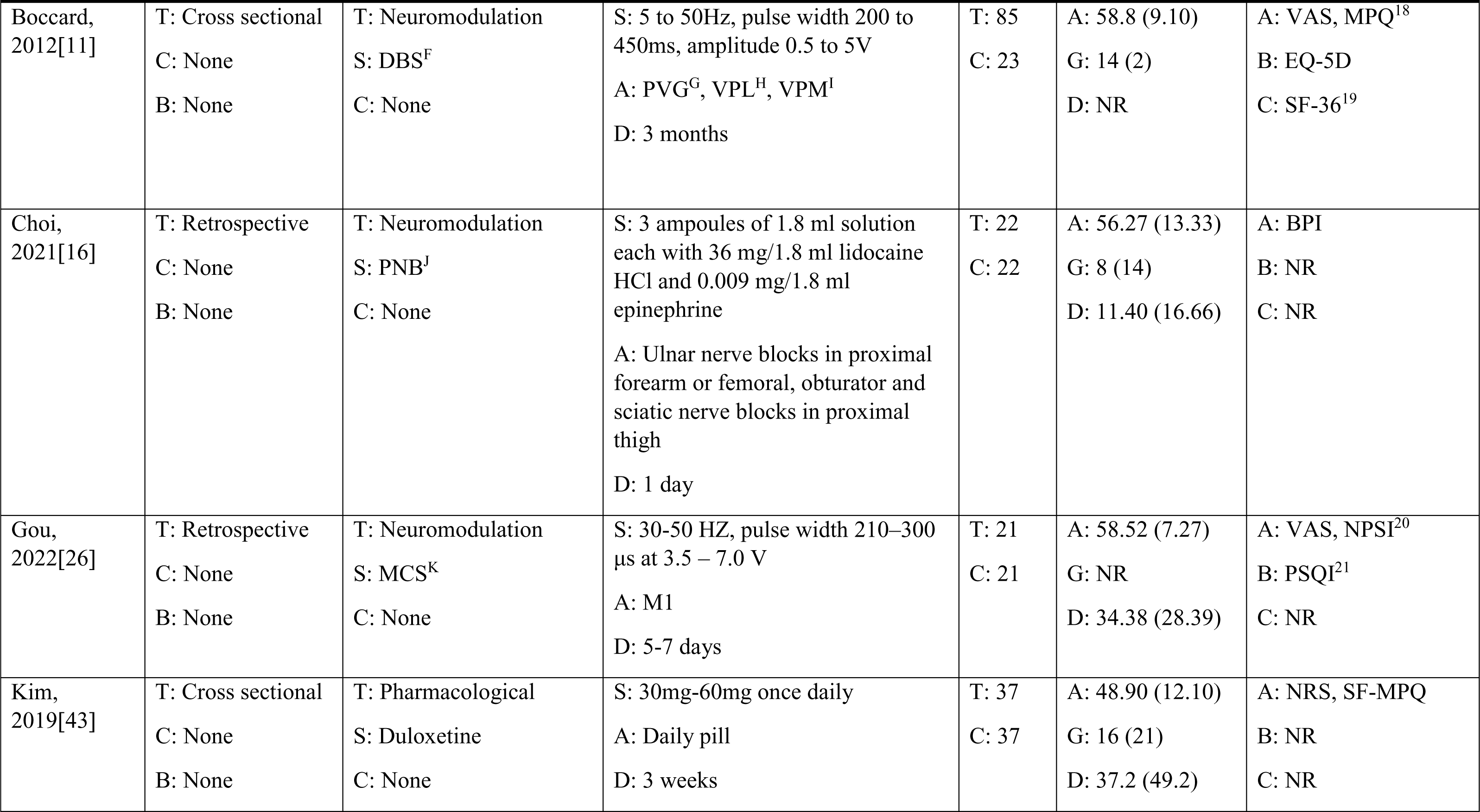
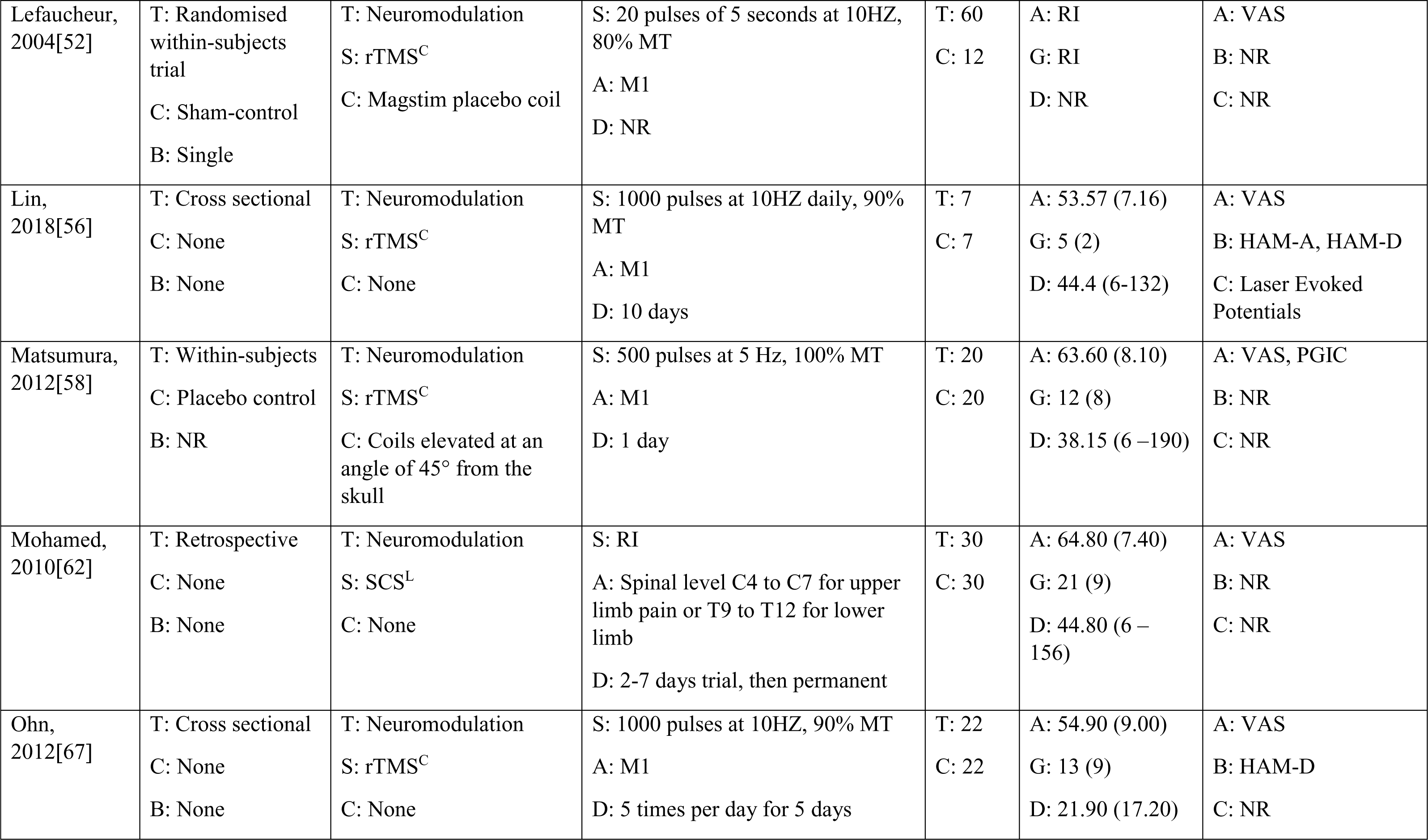
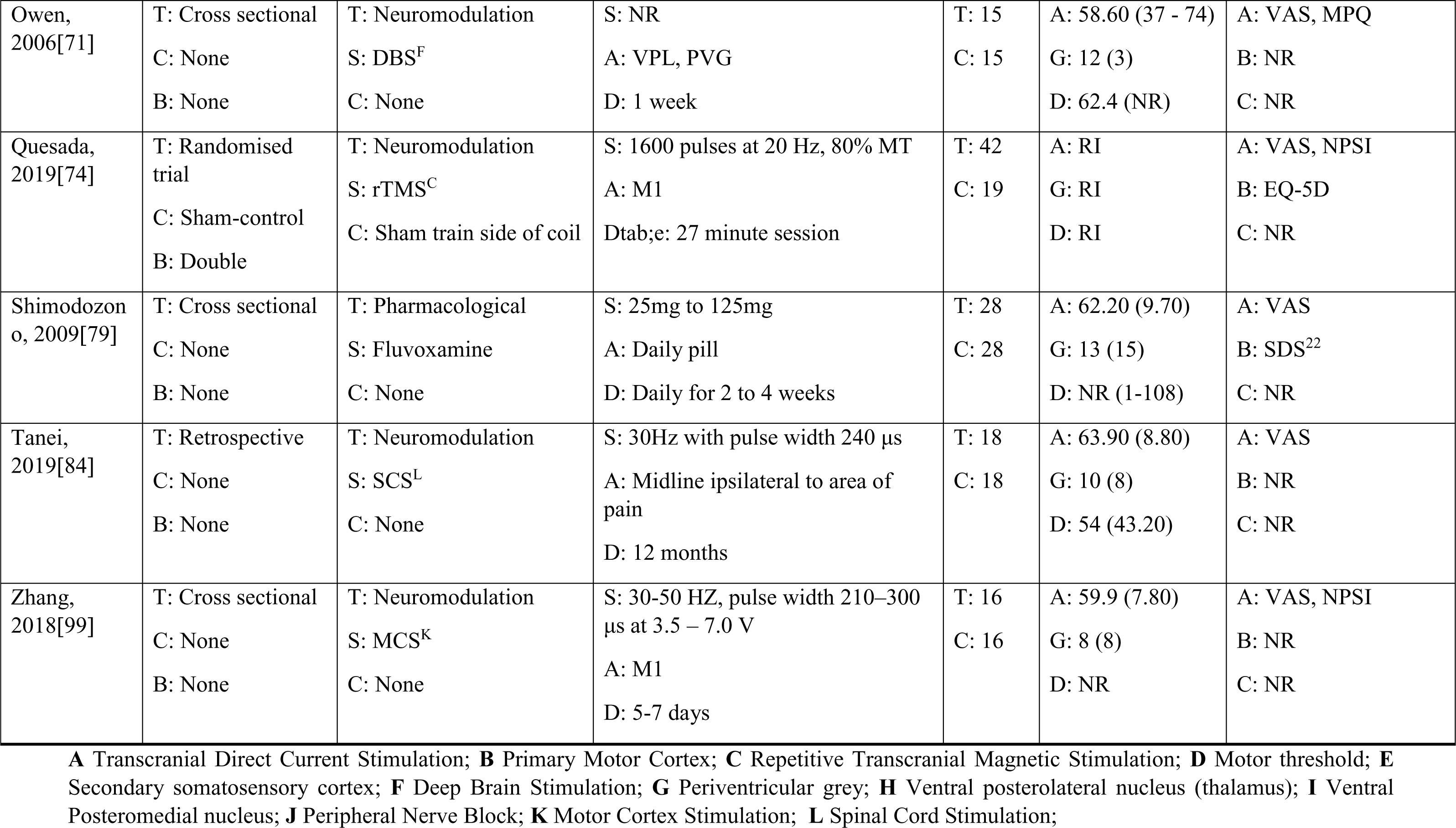
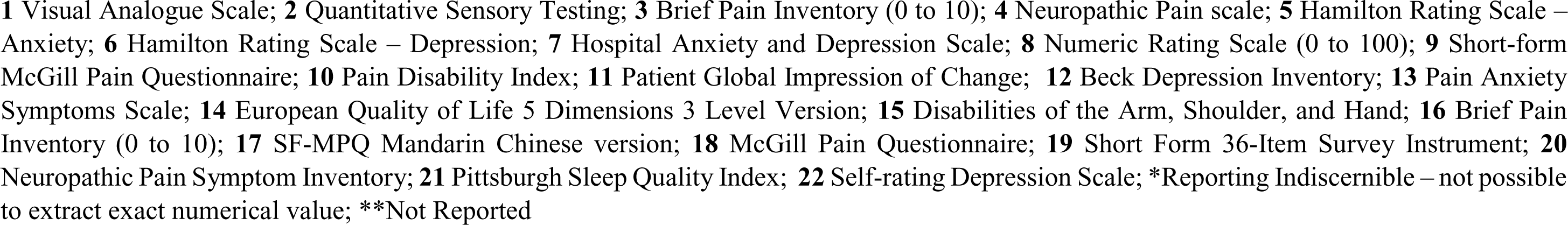
Study Characteristics; Random-Effects meta-analysis in grey, and proportional meta-analysis in white.

### 3.2 Risk of Bias and Certainty of Evidence

Risk of bias and certainty of evidence for individual studies included in either random-effects or proportional menta-analyses are presented in *Table 5*. The mean risk of bias rating for studies included in random-effects meta-analysis was “Some Concerns”, the mean risk rating for proportional meta-analysis studies was “Some Concerns”, and the mean risk rating for studies which were not included in meta-analyses was “Some Concern”. GRADE framework identified that random-effects meta-analysis included studies’ mean rating was “Moderate” certainty of evidence, proportional meta-analysis studies mean rating was “Low”, studies not included in meta-analyses had a mean rating of “Low”.

**Table 5.**
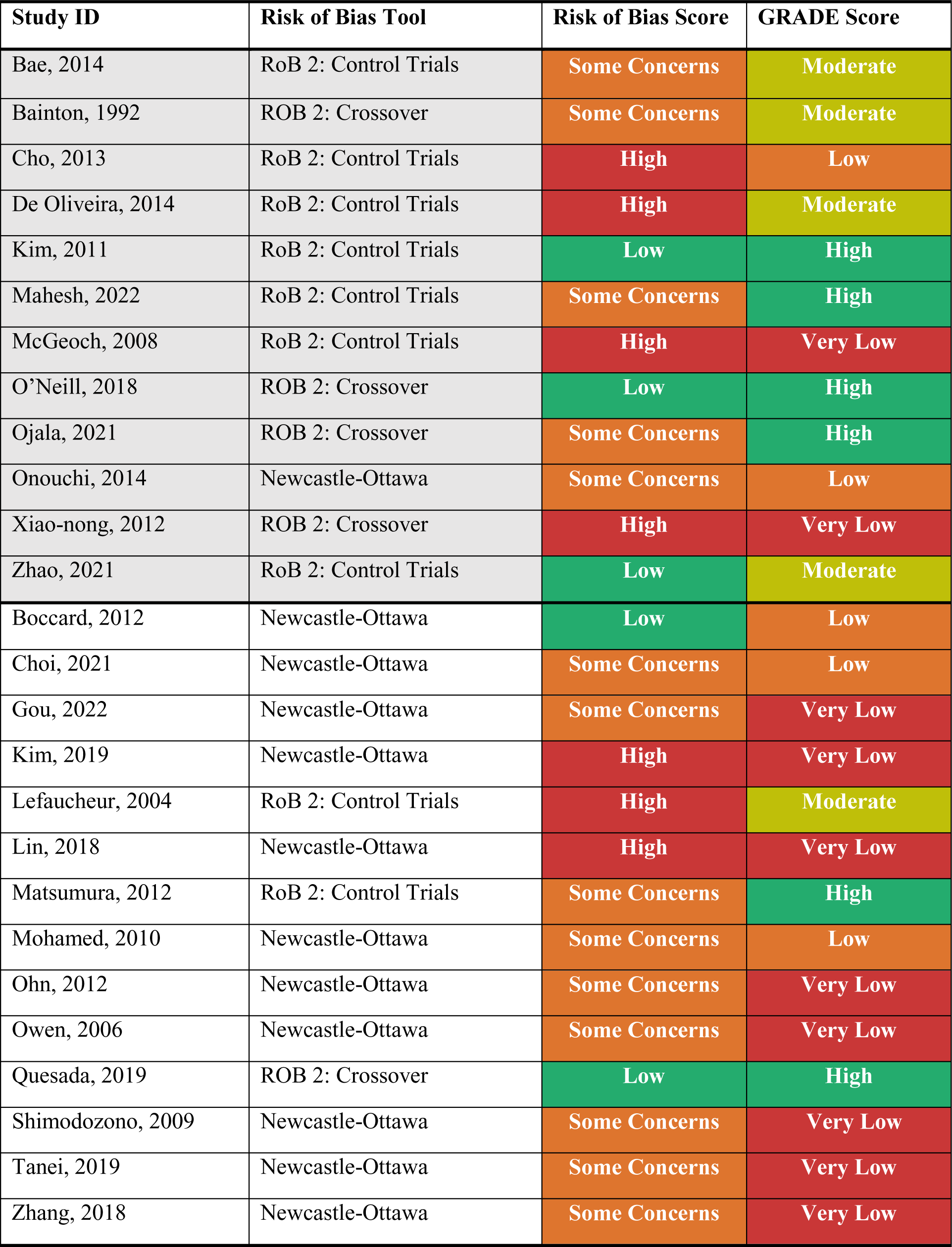
Study Risk of bias and Certainty of evidence; Random-Effects meta-analysis in grey, and proportional meta-analysis in white.

| Study ID | Risk of Bias Tool | Risk of Bias Score | GRADE Score |
| --- | --- | --- | --- |
| Bae, 2014 | RoB 2: Control Trials | Some Concerns | Moderate |
| Bainton, 1992 | ROB 2: Crossover | Some Concerns | Moderate |
| Cho, 2013 | RoB 2: Control Trials | High | Low |
| De Oliveira, 2014 | RoB 2: Control Trials | High | Moderate |
| Kim, 2011 | RoB 2: Control Trials | Low | High |
| Mahesh, 2022 | RoB 2: Control Trials | Some Concerns | High |
| McGeoch, 2008 | RoB 2: Control Trials | High | Very Low |
| O'Neill, 2018 | ROB 2: Crossover | Low | High |
| Ojala, 2021 | ROB 2: Crossover | Some Concerns | High |
| Onouchi, 2014 | Newcastle-Ottawa | Some Concerns | Low |
| Xiao-nong, 2012 | ROB 2: Crossover | High | Very Low |
| Zhao, 2021 | RoB 2: Control Trials | Low | Moderate |
| Boccard, 2012 | Newcastle-Ottawa | Low | Low |
| Choi, 2021 | Newcastle-Ottawa | Some Concerns | Low |
| Gou, 2022 | Newcastle-Ottawa | Some Concerns | Very Low |
| Kim, 2019 | Newcastle-Ottawa | High | Very Low |
| Lefaucheur, 2004 | RoB 2: Control Trials | High | Moderate |
| Lin, 2018 | Newcastle-Ottawa | High | Very Low |
| Matsumura, 2012 | RoB 2: Control Trials | Some Concerns | High |
| Mohamed, 2010 | Newcastle-Ottawa | Some Concerns | Low |
| Ohn, 2012 | Newcastle-Ottawa | Some Concerns | Very Low |
| Owen, 2006 | Newcastle-Ottawa | Some Concerns | Very Low |
| Quesada, 2019 | ROB 2: Crossover | Low | High |
| Shimodozono, 2009 | Newcastle-Ottawa | Some Concerns | Very Low |
| Tanei, 2019 | Newcastle-Ottawa | Some Concerns | Very Low |
| Zhang, 2018 | Newcastle-Ottawa | Some Concerns | Very Low |

### 3.3 Demographics of Included CPSP Participants

There were a total of 1451 participants included across 42 studies with 1191 of those being CPSP participants. The mean reported age of CPSP participants was 58.39 with a range of 23 to 82 years old. Out of the studies that reported CPSP participant gender, 58.76% (n = 597) were male and 41.24% (n = 419) were female. As only a small sample of studies reported participant race, this was omitted from data extraction.

### 3.4 Stroke Aetiology

Out of reported stroke aetiologies, 46.23% (n = 233) were ischemic and 53.77% (n = 271) were haemorrhagic. Most of the strokes were thalamic, making up 52.05% (n = 406) out of 780 reported cases. Extra-thalamic strokes made up 36.79% (n = 287), cortical strokes 8.85% (n = 69), and multimodal strokes 2.31% (n = 18) of reported cases. More specific distribution of stroke locations is illustrated in *Figure 2*, but the most common reported lesions were in the Thalamus, Brainstem and Putamen. Some studies did not specify the location of a stroke other than “Extra-Thalamic” or “Cortex”, and thus were labelled as “Undefined” (n = 86).

**Figure 2:**
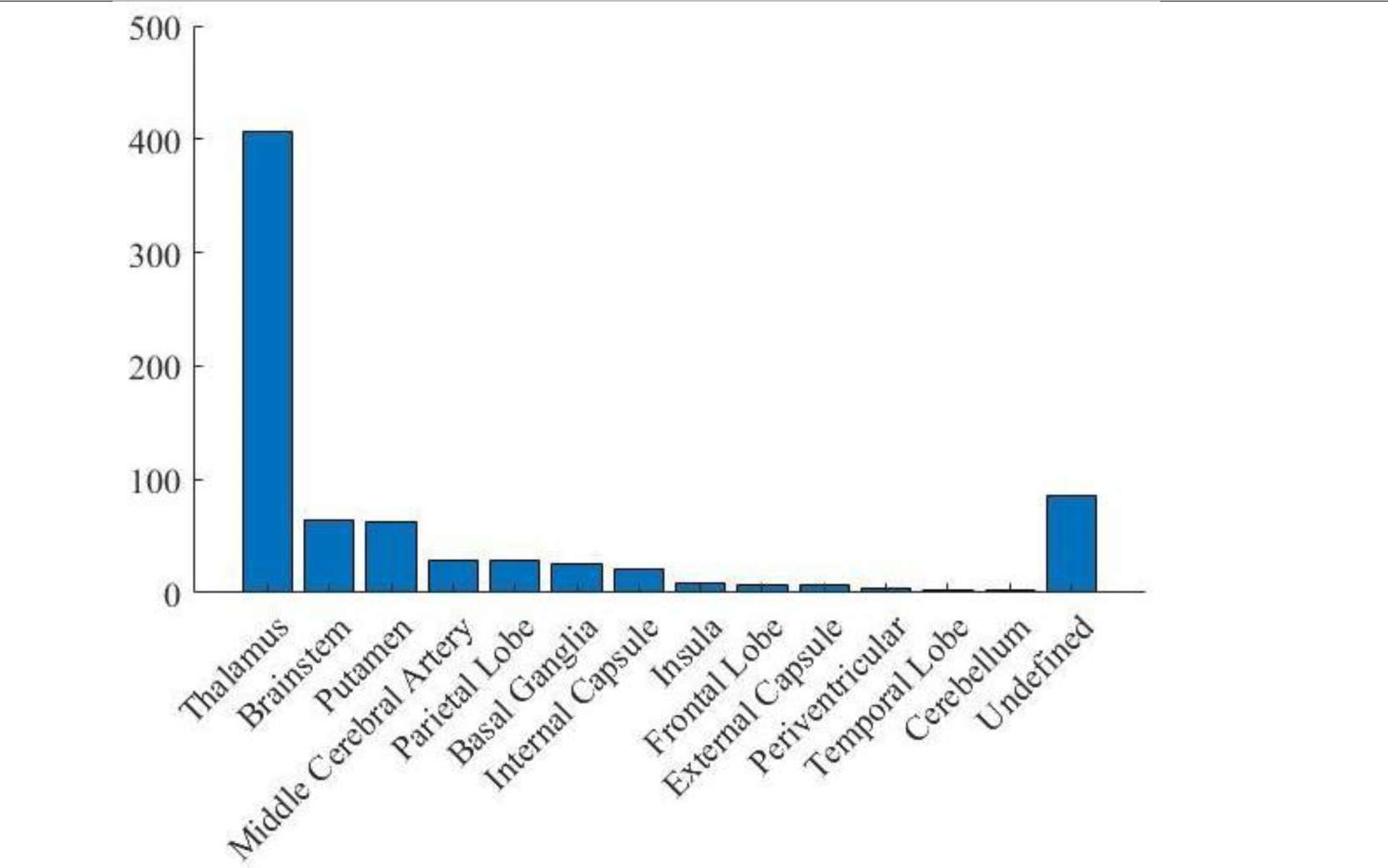
Distribution of CPSP participant stroke locations, y axis is number of participants, and x axis is the stroke location.

### 3.5 Specification of Pain

The mean duration of pain symptoms was 32.37 months with a range of 1 to 190 months across included studies. The mean pre-treatment VAS score across studies was 69.04. Pain was located on left side of the body in 52.05% of participants (n = 152), right side of the body in 47.26% of participant (n = 138), and in alternate sides in 0.69% (n = 2). Upper limb was the most common localization of pain with 30.92% (n = 175) of participants, hemibody pain was second most common with 30.39% (n = 172), lower limb pain was reported by 22.97% (n = 130), face localized pain by 12.36% (n=70), and trunk by 3.36% (n = 19) of participants. Regarding abnormal pain sensations, 43.52% (n = 245) of participants reported allodynia, 28.24% (n = 159) reported hyperalgesia, 11.19% (n = 63) reported spontaneous pain, 10.48% (n = 59) reported continuous pain, and 6.46% (n = 37) of participants reported hypoesthesia. The most common quality of pain reported was burning which was reported by 49.03% (n = 126) of participants, the second most common quality was tingling in 10.12% (n = 26), and third most common being tearing in 8.95% (n = 23) of participants. Full breakdown of reported qualities of pain is presented in *Figure 3*.

**Figure 3:**
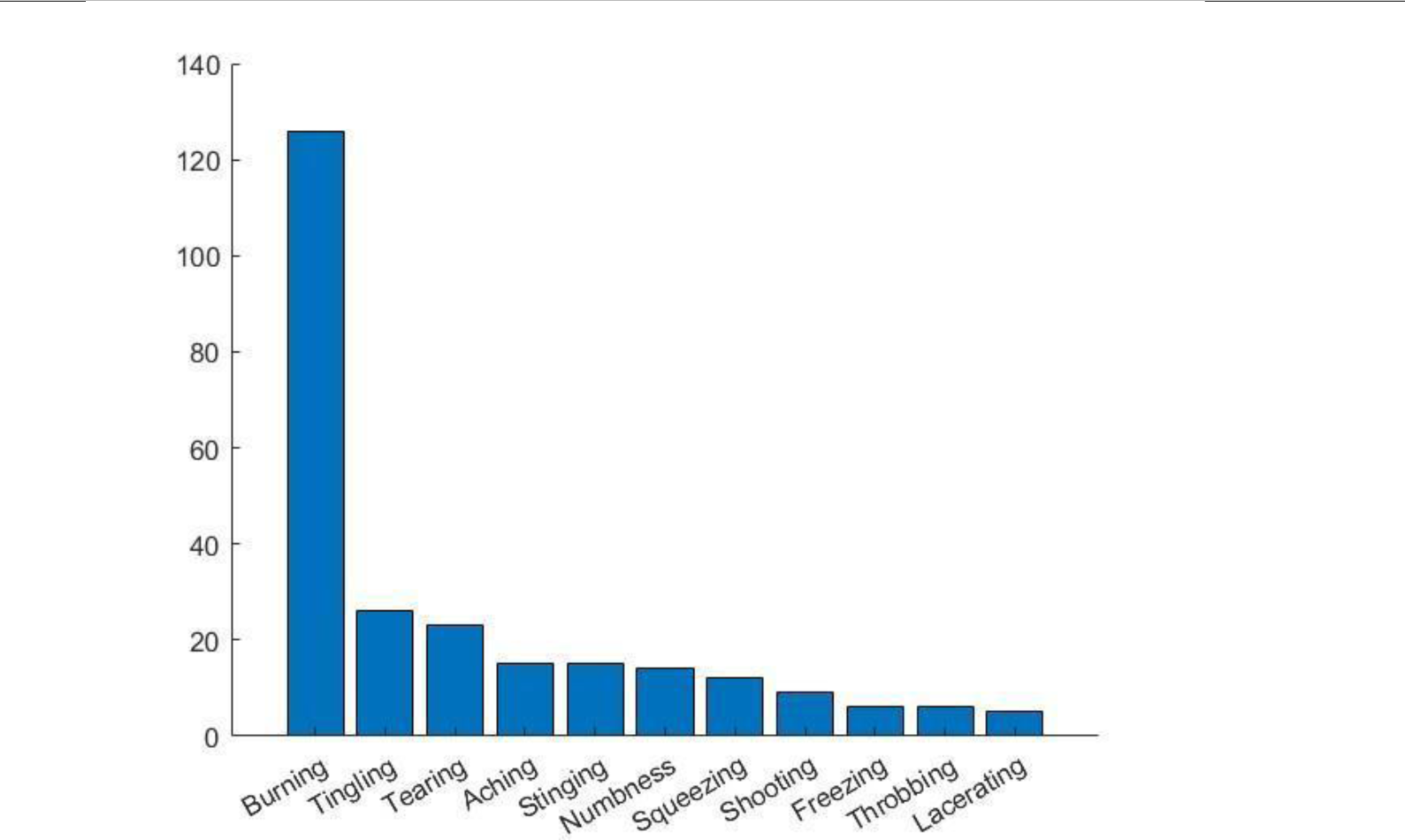
Distribution of CPSP participant qualities of pain, y axis is number of participants, and x axis is the pain quality.

### 3.6 Neuromodulation Interventions

#### 3.6.1 Random-Effects Meta-Analysis

Out of the five neuromodulation studies included in this analysis (n = 99), one rTMS study was split as per Cochrane guidelines into two groups – one for primary motor cortex and one for secondary sensory cortex. One other study used rTMS of primary motor cortex, one used rTMS of premotor cortex and prefrontal cortex, one used Transcranial Direct Current Stimulation (tDCS), and one used vestibular caloric stimulation. Across all neuromodulation studies, pain outcome was measured on average over 9.4 time points with final follow up being 0.48 months after last intervention. Neuromodulation was found to have a medium effect on mean VAS pain scores across these studies (SMD -0.60, 95% CI -0.97 to -0.23, I^2^ 7%; *Figure 4*). Funnel plot of studies showed symmetrical spread within 95% confidence, and Egger’s test of heterogeneity was not significant (p = 0.72). As this meta-analysis had low heterogeneity and moderate certainty of evidence, it would indicate that there is a consistent therapeutic effect of neuromodulation on CPSP.

**Figure 4:**
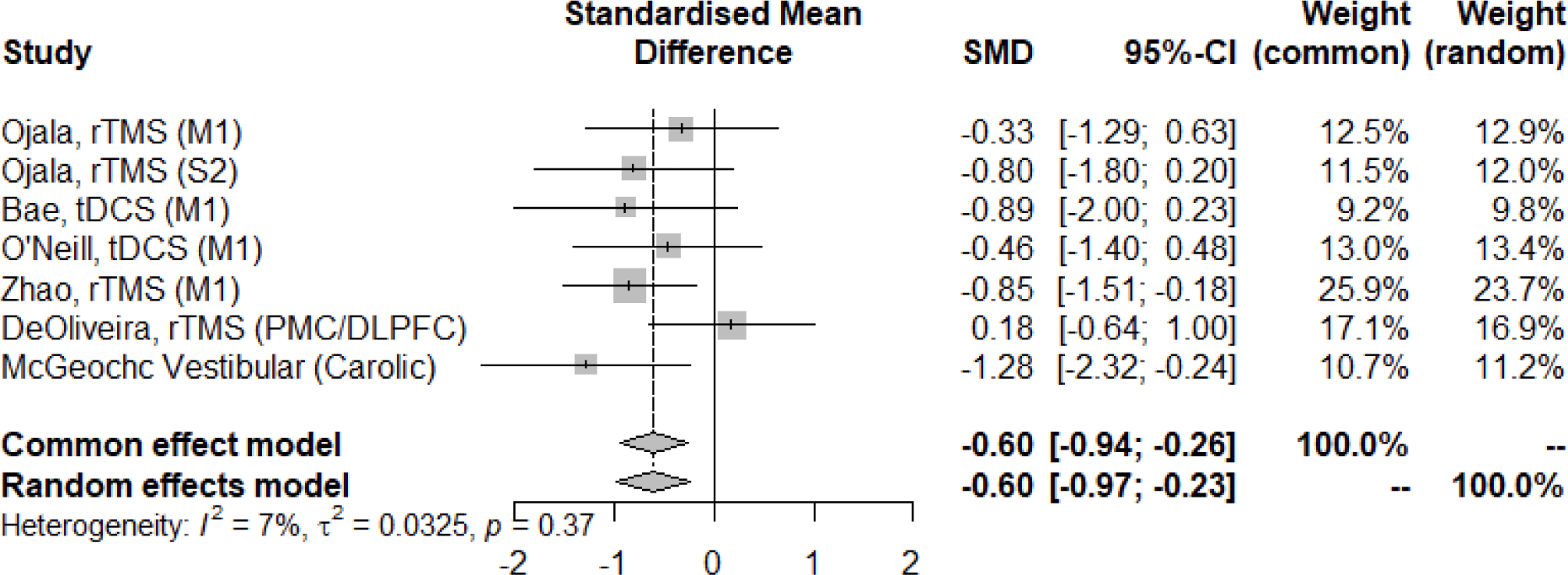
Meta-analysis results and forest plot from Neuromodulation studies.

A sub-group analysis of all rTMS studies (n = 76) found a small to medium effect size for mean VAS reduction (SMD -0.46, 95% CI -0.97 to 0.04, I^2^ 27.5%). Funnel plot showed symmetrical spread, and Egger’s test was not significant (p = 0.73). When only M1 targeting rTMS studies were included (n = 55), the effect size was medium (SMD -0.68, 95% CI -1.22 to -0.13, I^2^ 0%), but this analysis had high heterogeneity. Sub-group analysis of tDCS studies also found a medium effect size but high heterogeneity (SMD -0.64, 95% CI -1.35 to 0.008, I^2^ 0%).

#### 3.6.2 Proportional Meta-analysis

An analysis of 11 neuromodulation studies (n = 238), which included five rTMS, two Spinal Cord Stimulation (SCS), two Motor Cortex Stimulation (MCS), and two Deep Brain Stimulation (DBS) studies. On average, Pain outcome was measured over 6.09 time points with final follow up being 19.63 months after last intervention. The mean percentage reduction of mean VAS score across studies was 32.50% (CI 21.19 to 42.80, I^2^ = 99.13%). There was considerable and significant heterogeneity among the studies, which is illustrated by a forest plot in *Figure 5*.

**Figure 5:**
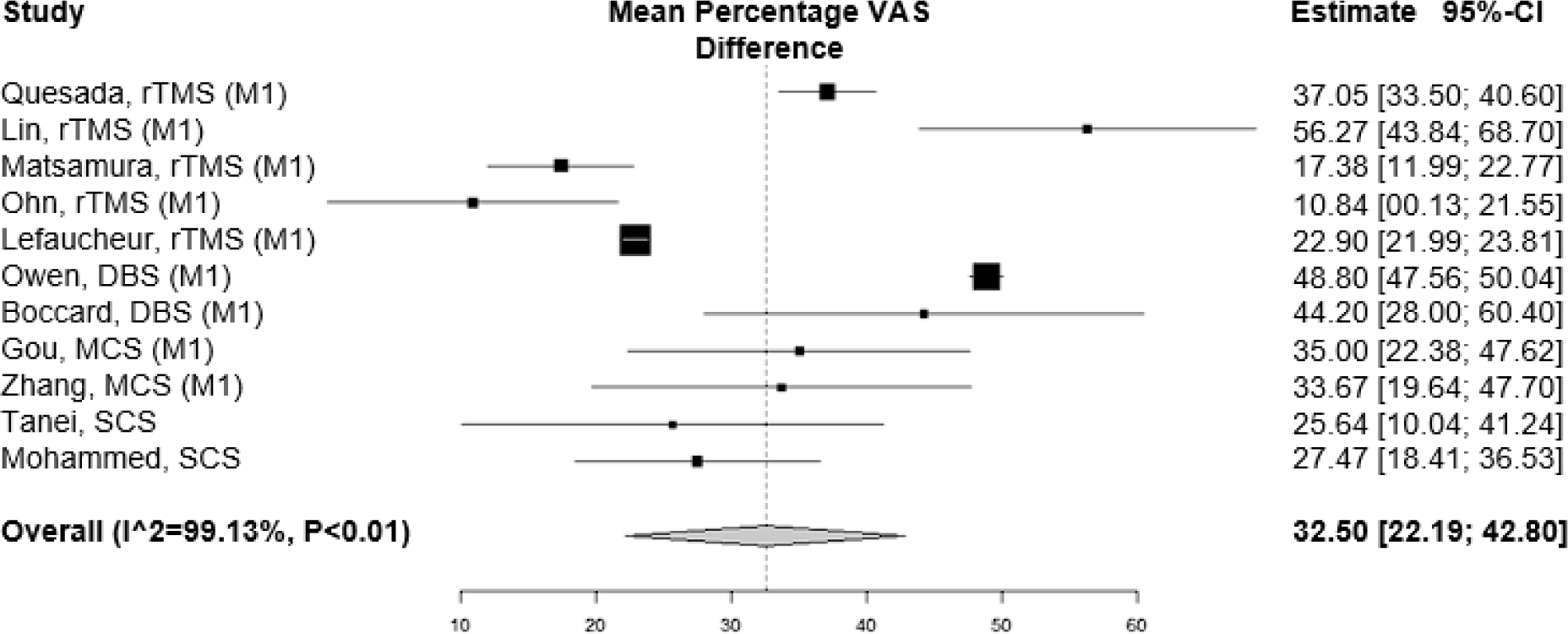
Proportional Meta-analysis results and forest plot from Neuromodulation studies.

A sub-group analysis of five rTMS studies found a reduction of mean VAS score by 26.50% with significant heterogeneity (CI 17.86 to 35.12, I^2^ = 95.05%). Sub-group analysis of two M1 DBS studies found a reduction of mean VAS score by 48.77% with low heterogeneity (CI 47.53 to 50.01, I^2^ = 0%). Sub-group analysis of two MCS studies found a reduction of mean VAS score by 34.90% with low heterogeneity (CI 25.52 to 44.28, I^2^ = 0%). Sub-group analysis of two SCS studies found a reduction of mean VAS score by 27.00% with low heterogeneity (CI 19.17 to 34.84, I^2^ = 0%).

#### 3.6.3 Narrative Summary

There were four rTMS studies which were not included in either meta-analysis. Across these studies, outcomes were measured, on average, over 10.25 time points with final follow up being 0.46 months after last intervention. Khedr[41] is a placebo-controlled trial of rTMS over the primary motor cortex, but was not included in meta-analyses due to not reporting CPSP specific VAS reduction (n = 24), and instead reported all sample reduction (n = 48). It was reported that 50% of all neuropathic pain patients achieved satisfactory (over 40%) VAS reduction 2 weeks after last session and 14.3% achieved good (over 70%) pain reduction. This was compared to the control group where 100% of neuropathic pain participants did not achieve satisfactory pain control. Kobayashi[46] performed a cross-sectional within subjects sham-controlled single blind study of rTMS over the motor cortex. Out of their 18 participants, 13 showed VAS percentage reduction of over 40% with 5 of those patients achieving over 79% reduction. The mean VAS reduction from 8^th^ week of treatment to 12^th^ weeks was 36.7 (SD = 21.1, p <0.01). McLean[60] performed a cross sectional rTMS study of 50 neuropathic pain participants, of which 5 had CPSP. After 6 weeks of stimulation CPSP participants reported a 53 point mean pain score decrease, but significance was not reported nor SD or metrics to impute SD were provided. The final study[27] performed a cross-sectional study of rTMS over primary motor cortex without a sham control (n = 14). The mean VAS score reduction was 7.00 or 10% on a 101 point scale (p = 0.02). However, after 4 weeks the score increased by 3 points and was only a 3-point decrease from baseline and no longer significant (p = 0.17). Taking all rTMS studies into account, 66.07% (n = 37) out of 56 participants achieved satisfactory pain control as measured by over 40% VAS reduction.

There were three cortex stimulation studies not included in the meta-analyses. Tsubokawa[86] performed a cross-sectional study without a control or sham group. Across these studies, outcomes were measured, on average, over 65.33 time points with final follow up being 20 months after last intervention. Out of 11 participants, 9 achieved pain reduction of over 50% and 6 of those achieved pain reduction of over 80%. Longitudinal results showed that 45% of participants achieved pain control (p < 0.05) over 2 months which is broken down to 5 patients maintaining over 80% of pain reduction, 1 participant having between 40% and 60% reduction, and 5 participants having less than 40% pain reduction. However, it was reported that tolerance to treatment developed within 7 months and good pain control reduced to only 38% of participants after 2 years. Yamamoto[97] study utilised cross sectional within-subjects comparative study of motor cortex stimulation versus pharmacological treatment. Monopolar square wave pulses were delivered to the motor cortex of 39 participants several times per day. Out of those participants, 13 were reported to have reduced pain by over 40% which was statistically significant (p < 0.05). The final motor cortex stimulation study[40] investigated the effect of chronic motor cortex stimulation of 31 participants in a cross-sectional study without sham or control. Participants were tested for 1 week before implantation and 23 participants saw pain being reduced by over 40%. After implantation, 15 out of 23 participants maintained pain reduction of over 40%. Significance metric was not reported for these reductions. Overall, from 81 participants across MCS studies, 41.98% (n = 34) achieved satisfactory pain control.

Yamamoto[98] performed a cross sectional within-subjects comparative study of spinal cord stimulation effectiveness versus pharmacological treatment. VAS mean pain was measured over 12 time points, with the last being 5 minutes after last injection. Of the 22 participants, 15 received VAS score alleviation of over 30%, and 6 of those received pain alleviation of over 60%. Significance was not reported for alleviation, but it significantly added to pain alleviation for ketamine treatment as add-on therapy. No side effects or other improvements were reported.

### 3.7 Secondary Outcomes and Side Effects

Out of all rTMS studies, Neuropathic Pain Inventory, Hamilton Depression Rating Scale (HAM-D) and Hamilton Anxiety Rating Scale (HAM-A) scores showed no significant improvement. Two rTMS studies[49,58] reported Patient Global Impression of Change (PGIC) improvement in 17 out of 34 participants across these studies. From 12 rTMS studies, 7 reported side effects and across those studies, 16% (n = 32) out of 200 reported mild adverse side effects, which included: increased pain, slight scalp discomfort, headache, dizziness, tiredness, paraesthesia, and facial muscle twitching. The most common side effects were transient increased pain (n = 6), scalp discomfort (n = 5), and headaches (n = 4). There was 1 moderate adverse event which was collapse.

The one study on tDCS[6] reported that there were no significant differences in either cold sensation, warm sensation, cold pain or heat pain thresholds. No side effects were reported.

Vestibular Stimulation study[59] did not report any secondary outcome improvements or side effects. Two SCS studies[62,98] did not report any secondary outcomes, but did report an interaction of age and stroke location – younger participants and participants with thalamic stroke were more likely to respond to treatment. No side effects were reported for either study.

From all 5 MCS studies, only one study reported secondary outcomes[26]. They found that MCS significantly improved Pittsburgh Sleep Quality Index score from mean 16.38 score to mean 13.95 score. Side effects were reported in 3 MCS studies[40,86,97], and mild adverse events were seen in 3.70% (n = 3) out of 81 participants: 1 participant had a postoperative infection, 2 participants reported increased pain. There were 3 patients (3.70%) with generalised seizures when high-frequency pulses went above muscle contraction threshold.

### 3.7 Pharmacological Interventions

#### 3.7.1 Random-Effects Meta-Analysis

There were four pharmacological studies included in the random-effects meta-analysis (n = 231): two anticonvulsant (Pregabalin) studies, one antidepressant (duloxetine) study, and one opioid (Naloxone) study. On average, Pain outcome was measured over 3.5 time points over 15.53 months of intervention but long-term follow up was not carried out in any study. Pharmacological interventions were found to have a small effect on mean VAS score (SMD - 0.36, 95% CI -0.68 to -0.03, I^2^ 60%; *Figure 6*). Funnel plot showed symmetrical spread within 95% CI, and Egger’s test of heterogeneity was not significant (p = 0.60). Overall risk of bias score was “Some Concerns” and certainty of evidence was moderate to high, which indicates that there is a consistent therapeutic effect. A sub-group analysis of two anticonvulsant studies found a smaller effect on VAS reduction than overall pharmacological intervention (SMD - 0.31, 95% CI -0.78 to 0.15, I^2^ 72.5%). This meta-analysis indicates considerable heterogeneity, contains only two studies and moderate certainty of evidence, so this effect is limited in interpretation.

**Figure 6:**
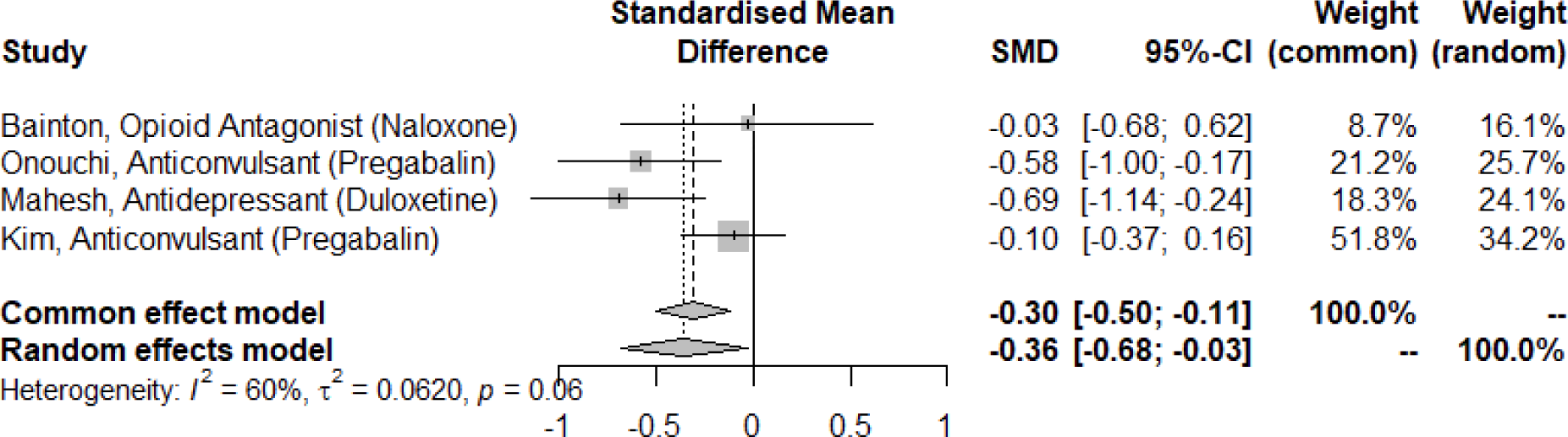
Random-Effects Meta-analysis results and forest plot from Pharmacological studies.

#### 3.7.2 Proportional Meta-analysis

The analysis was conducted on three pharmacological studies (N = 71): two antidepressant studies and one study of Lidocaine delivered as Peripheral Nerve Block (PNB). Results showed that the total percentage reduction of mean VAS score was 58.33% (CI 36.51 to 80.15, I^2^ = 84.23%). There was considerable and significant heterogeneity among the studies, which is illustrated by a forest plot in *Figure 7*. Across these studies, pain outcomes were measured over 2 time points – before and after interventions – over, on average, 1.02 months of treatment. Choi^15^, was the only study with a long-term follow up at 1 month, which was the score used to calculate percentage pain reduction. Sub-group analysis of two antidepressant studies found a reduction of mean VAS score by 67.43% with significant considerable heterogeneity (CI 26.85 to 108.02, I^2^ = 91.25%).

**Figure 7:**
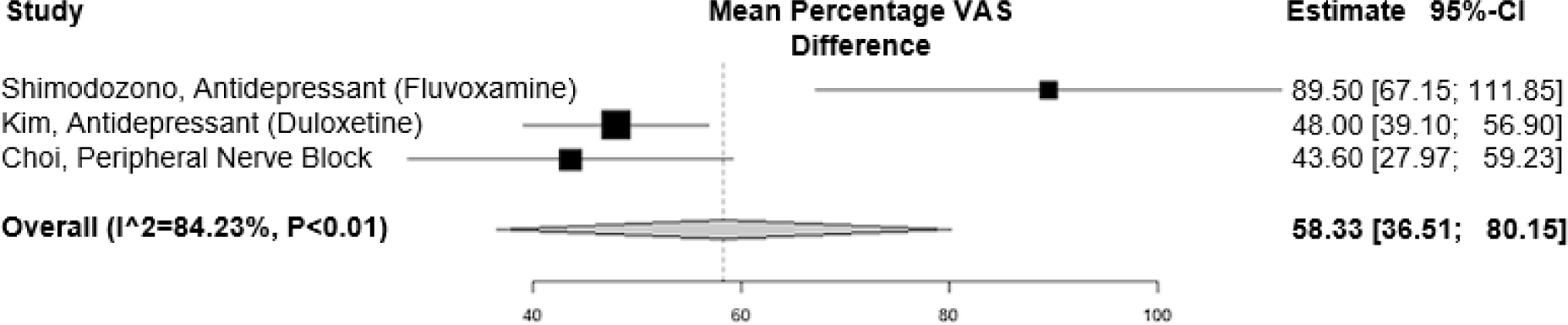
Proportional Meta-analysis results and forest plot from Pharmacological studies.

#### 3.7.3 Narrative Summary

There were two antidepressant studies which were not included in either meta-analysis due to not reporting SD metrics. Lampl[50] performed a randomised placebo-controlled double-blind trial of 39 participants treated with amitriptyline as prophylaxis to CPSP. After 3 weeks of titration of up to 75mg of Amitriptyline, 1 less participant developed CPSP in Amitriptyline group than the placebo group and the mean VAS score was 2.0 smaller in the intervention group. These findings were not significant. Leijon and Boivie[53] performed a randomised placebo-controlled crossover study of 15 participants treated with 75mg amitriptyline (antidepressant) and 800mg carbamazepine (anticonvulsant) on mean pain score. Each medication period had 28 VAS assessment time-points, but no longitudinal follow-up. After 4 weeks, only amitriptyline had a sustained significant reduction of pain with 10 patients reporting reduced pain with a mean 5 point reduction from initial mean 47 point score.

There were five more anticonvulsant studies which could not be included in meta-analyses. Across these studies, pain assessments were taken, on average, over 14.4 time points over a treatment period of 2.95 months. No study measured longitudinal follow up scores. Jungehulsing[38] conducted a randomised placebo crossover double-blind trial of 42 participants for levetiracetam as CPSP treatment. The mean levetiracetam dose was 2130mg but it did not have a significant effect on participant mean pain scores. Rahajeng[76] performed an observational study of 36 participants taking pregabalin for CPSP. After 12 weeks, 26 participants had more than 26% mean pain reduction, which was a significant effect (p < 0.05), and 6 of those participants had over 51% pain reduction. Kalita[39] investigated pregabalin and lamotrigine in a crossover 30 participant open-label design. The maximum dose of pregabalin at the end of 8 weeks was 600mg daily and lamotrigine dose was 200mg daily. Out of those participants, pain alleviation of over 50% was achieved by 19 participants with pregabalin (p < 0.0001) and 16 with lamotrigine (p < 0.0001). These studies would indicate that out of 66 participants across both Pregabalin studies, over 50% pain reduction was achieved in 37.88% (n = 25) of participants. Petramfar[73] performed a retrospective review of 17 participants taking lamotrigine for CPSP. After 24 weeks, there was a significant mean pain score decrease by 24.1 points (p = 0.001) from initial mean score of 68.20. Vestergaard[91] conducted a randomised placebo-controlled crossover double-blind trial of 30 participants for lamotrigine as treatment for CPSP. The median initial pain score was 60 and the median pain score after treatment was reduced by -10 compared to placebo of increase by 10. This was a significant reduction in pain scores (p = 0.02), but the decision to report median rather than mean score was not described or justified.

There was one anaesthetic study[68] which investigated the effectiveness of morphine, thiamylal and ketamine as compared to MCS and placebo saline. This was a cross sectional, within subjects design with 39 participants. Anaesthetics were titrated on different one-day sessions with washout periods. Morphine was injected up to 18mg, thiamylal was injected up to 250mg, and ketamine was injected up to 25mg. Effectiveness of each treatment was measured by the number of patients with reduced pain, which were: 8 for morphine, 22 for thiamylal, 11 for ketamine, and 13 for MCS. The percentage or mean reduction of pain was not reported.

#### 3.7.4 Secondary Outcomes and Side Effects

A study of duloxetine intervention[43] was the only one to not report secondary outcomes. On the other hand, Mahesh[57] duloxetine study found a significantly increased PGIC score when compared to the placebo group. From 119 participants across both studies, side effects were seen in 36.13% (n = 43). Reported symptoms included: nausea, agitation, somnolence, dizziness, and recurring vomiting. Studies of amitriptyline and carbamazepine[50,53] did not have a significant effect on depression scores. Across both studies, 64.81% (n = 35) out of 54 total participants reported mild reactions, which included tiredness, dry mouth, vertigo, and gait disturbances. Further 3.70% (n = 2) reported undisclosed moderate side effects. Fluvoxamine was found to have a significant effect on the Self-rating Depression Scale with a 7.7 point decrease (p < 0.01) from initial 44.3. Out of 28 participants, 10.51% (n = 3) withdrew due to side effects, but side effects of the remaining participants were not tracked. Across all antidepressant studies, 36.82% (n = 74) out of 201 participants reported side effects.

Levetiracetam was not found to have a significant effect on McGill Pain Questionnaire or Beck Depression Inventory. There were 34 counts of mild adverse events reported in levetiracetam intervention group: 11 reported tiredness, 8 reported pain increase, 7 reported dizziness, 4 reported pruritus, and 4 reported headaches. There were 7 people who reported withdrawals from levetiracetam with symptoms of fatigue and pain increase. Pregabalin was found to significantly improve allodynia, HAM-A and sleep However, out of 378 pregabalin participants across both studies[39,42], side effects were reported in 56.35% (n = 213). These included: somnolence, tremor, sedation, dizziness, pedal oedema, peripheral oedema, blurred vision, weight gain, and irritability. Out of all side effects, 10 were moderate and 8 were severe. Withdrawal was not observed or recorded in any of these studies. Across 2 studies which reported other improvements with lamotrigine treatment, it was found that HADS score, allodynia, sleep, and mood significantly improved. From the 3 lamotrigine studies[39,73,77,90], mild side effects were reported in 36.36% (n = 28) and moderate to severe side effects were reported in 7.79% (n = 6) out of 77 participants. These side effects included: skin rash, somnolence, dizziness, fatigue, nausea, severe headaches, and severe pain. Overall, anticonvulsant studies found 54.29% (n = 247) out of 455 participants experienced side effects.

From the study of anaesthetics[68], no secondary improvements were reported. Out of all treatments, ketamine was the only drug to induce side effects. 5.13% (n = 2) out of 39 people experienced transient abnormal sensations. From a study of opioid antagonist Naloxone[7], no secondary outcomes were reported. There were 3 participants (15%) out of 20 who reported adverse reactions: 1 with increased pain and 2 with substantial rise in pulse due to sweating, tremor, salivation and pain.

A peripheral nerve block study[16] reported that patients who responded to PNB showed a significantly reduced impact of pain on ability to work, relations with other people, and non-significant improvements of mood (p = 0.10) and sleep (p = 0.06). No participants reported adverse effects other than some transient weakness and numbness that lasted up to 3 hours.

### 3.8 Physical Interventions

#### 3.8.1 Random-Effects Meta-Analysis

An analysis of two physical acupuncture studies found to have a moderate effect on mean VAS score (SMD -0.55, 95% CI -1.28 to 0.18, I^2^ 0%; *Figure 8*). Both studies only collected VAS scores once before and once after intervention over a mean of 0.39 months. Funnel plot showed symmetrical spread within 95% CI, but heterogeneity could not be reliably tested with 2 studies. Overall risk of bias score was “High” and certainty of evidence was low to very low, which severely limits the interpretation of this analysis.

**Figure 8:**
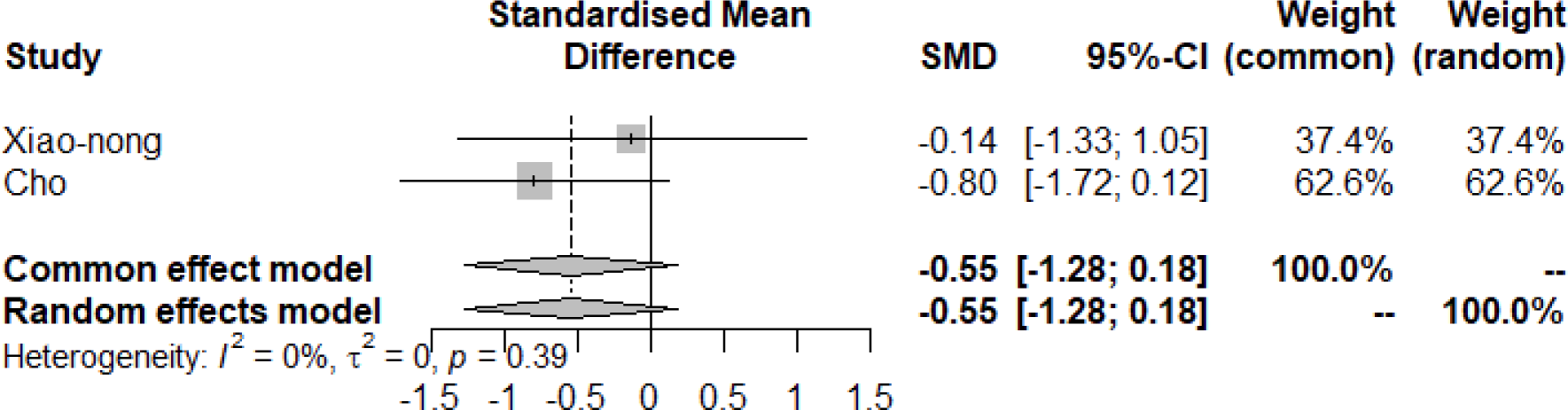
Random-Effects Meta-analysis results and forest plot from Acupuncture studies.

#### 3.8.2 Narrative Summary

No physical intervention studies were included in the proportional meta-analysis. Simmonds and Shahrban[82] cross sectional sham controlled within subjects study exposed 12 participants to either “hot world”, “snow world”, neutral stimuli, or no stimuli. “Hot world” included scenes of volcanoes, “snow world” included snowy mountains, and neutral stimuli consisted of alternating white pillars on a black background. “Hot” stimuli significantly decreased participant mean VAS score by 17 points on a 101 point scale (p = 0.01), “Cold” stimuli significantly decreased it by 17 points (p = 0.01), neutral stimuli showed a 3.8 point increase (p = 0.87), and no VR showed a 3.0 point increase. Pain scores were collected only twice – before and after intervention – on the same day. Other improvements included significant increased threshold to cold and heat stimuli. No side effects were reported.

#### 3.8.3 Secondary Outcomes and Side Effects

Neither of the two acupuncture studies reported any secondary improvements[15,95]. Across both studies, only 1 person (3.70%) out of 27 experienced side effects from acupuncture who left the study due to itching. Virtual reality[82] was found to significantly increase threshold to cold and heat stimuli. No side effects were reported.

### 4.1 Discussion

This systematic review and meta-analysis set out to review 4 different intervention types: pharmacological, physical, psychological, and neuromodulation. However, no psychological intervention studies for CPSP patients were found in this review. There were 15 pharmacological, 3 physical, and 24 neuromodulation intervention studies. Stroke lesion causes were about equally distributed between ischemic and haemorrhagic. The most common reported stroke lesion location was thalamus, with brainstem and putamen as second most common locations, but this may be due to older studies focusing on thalamic origin pain. Duration of CPSP varied greatly from 1 month to 190 months, but the average was around 32.57 months. Time from stroke until CPSP onset was rarely reported across studies. Lateralization of pain was evenly divided between the right and left side of the body. Hemibody, upper limb, lower limb, and face were the most common areas of pain. Allodynia was the most commonly reported pain characterization, followed by hyperalgesia and spontaneous pain. Mean VAS score before treatment for CPSP participants was 69.04, with most patients reporting the pain as “burning”, which was the most common descriptor and used almost 5 times more than the next most common descriptors: “tingling” or “tearing”.

From investigation of random-effects meta-analyses, neuromodulation was the most effective treatment with a moderate effect size, followed by physical interventions with moderate effect size, and pharmacological interventions with a small effect size. However, physical studies were limited by high risk of bias and low level of certainty^15,82,95^. Sub-group analysis of rTMS and tDCS found moderate effect sizes, whereas anticonvulsants were found to have a small effect size. The rTMS effect size was limited by one study^5^ which applied rTMS to the premotor cortex and did not find a significant effect. Additionally, analysis of only rTMS interventions targeting M1 found the effect size to be higher than overall neuromodulation effect and slightly higher than tDCS, which supports the use of rTMS and tDCS as promising alternatives to invasive neuromodulation treatments[27,80,96]. The therapeutic action of modulating M1 is still being investigated, but could relate to inadvertent signalling of primary sensorimotor cortex (S1) or the potential pain pathway between M1 and S1[20]. Previous review articles[21,55,94,96] have pointed out a lack of standardised protocol for rTMS, but majority of the studies presented in this review adhere to 10-20Hz at 80-90% of motor threshold. There is not enough data to compare effectiveness at different frequencies, but future work is needed to clearly define the most optimal stimulation parameters.

Proportional meta-analyses were only performed on pharmacological and neuromodulation studies, as there were not enough physical studies for inclusion. Pharmacological interventions were found to be more effective than neuromodulation studies. However, significant heterogeneity of both meta-analyses reduces generalisability. Sub-group analysis found that antidepressants and DBS were the most effective in reducing VAS pain percentage in pharmacological and neuromodulation groups respectively. Conversely, a separate review of pharmacological CPSP treatments found gabapentin and pregabalin to be more effective[13], but the present review included a wider literature pool while some studies cited in Chen and Li[77] review could not be accessed in English. No other review articles have reliably compared pharmacological CPSP interventions, but animal research supports antidepressants as the most effective pharmacological treatment[81].

From narrative analyses, comparison of rTMS and MCS found that rTMS studies had more participants achieve satisfactory pain control at over 40%. The one SCS study[98] used 30% as a satisfactory control benchmark and, thus, cannot be effectively compared with rTMS or MCS. There was only one prophylactic study which found that Amitriptyline is not effective in preventing CPSP[50], but another study indicates that it could be used to reduce pain scores in CPSP patients[53]. Pregabalin was the most effective anticonvulsant in reducing mean pain score, followed by Lamotrigine, and Levetiracetam which had a non-significant effect. These findings are in line with findings from a recent post-stroke pain pharmacotherapies review which identified Pregabalin as the most effective treatment for CPSP[10]. A comparison of anaesthetic medication found that thiamylal was the most effective treatment when compared with ketamine or morphine. Only one physical study was identified[82] - a VR trial that demonstrated, immediately following treatment, significantly reduced mean VAS pain score and increased pain threshold for cold and heat stimuli. VR has been used in some capacity for post-stroke rehabilitation and chronic pain management, but is rarely used on CPSP patients[1,51,54]. Further research is needed to reliably understand VR effectiveness for CPSP, but it should be explored as a potential non-pharmacological and non-invasive add-on treatment.

Secondary outcomes were only reported in select studies. Mood, anxiety or depression scores were not improved by rTMS, amitriptyline or carbamazepine. Amitriptyline[50] was only given for 3 weeks, which may not have been enough time to provide full effects, while the carbamazepine trial[53] reported that participants had low baseline depression scores. Fluvoxamine[79], Pregabalin[39,42,76], Lamotrigine[73], and MCS[26] all resulted in improvements in wellbeing through selective improvement of sleep, anxiety and depression scores. Peripheral nerve block had a significant effect on ability to work, but it is likely that this is due to a reduced pain score rather than the treatment itself. PGIC was only reported as significant in rTMS[27] and duloxetine studies[57], which might be confounded by the experience of rTMS, and mood stabilising properties of duloxetine. Improvement of allodynia was only reported in Lamotrigine[73], Pregabalin[39] and VR[82] interventions, but allodynia was rarely tested or reported across studies and further research is needed to understand how other interventions impact this symptom. Pregabalin and Lamotrigine effectiveness has been evidenced in literature, but VR is rarely explored for allodynia treatment[54,85,90]. Although, findings from this VR study would support the connection of VR as a tool to help control emotional feedback to pain which could moderate pain perception[88].

Virtual reality was the only intervention which had no side effects reported by participants. Acupuncture[15,95] and anaesthetics[97,98] had second least side effects reported, but these studies were limited by high risk of bias, low certainty of evidence, and small sample sizes. Acupuncture studies[15,95] were exclusively performed in East Asian countries where use and belief in acupuncture benefits are more widely accepted, which limits generalisation for populations in other countries. From least to most side effects reported interventions were: MCS, naloxone, rTMS, antidepressants, and anticonvulsants. MCS had the least amount of side effects measured over longest and most frequent follow-up times which indicates longitudinal pain relief, but it is an invasive and intensive procedure that should not be considered as primary intervention[55]. Antidepressants and rTMS were the only treatments with moderate adverse reactions, but they did not report any serious reactions. Anticonvulsant studies had the most reported side effects, but they had by far the largest sample which likely influenced side effect incidence. Anticonvulsant studies were also the only ones which had participants report withdrawal symptoms.

All things considered, antidepressants appear to be the most accessible treatment, particularly fluvoxamine, with a moderate effect on pain reduction, reported improvement in PGIC and mood, and least incidences of side effects and withdrawals. Anticonvulsants, particularly Pregabalin, could be an alternative if antidepressants are not well tolerated or effective. This is followed by rTMS and tDCS as effective non-invasive neuromodulation interventions. Differences between rTMS and tDCS effectiveness for neuropathic pain has been explored in literature with findings suggesting close to equal performance with some preference for the former, but responders for either modality do not tend to overlap so both treatments should be trialled for pain alleviation[23,33,42]. Moreover, a recent review has suggested rTMS is a more advantageous treatment than pharmacological interventions due to a single-target mechanism that does not disrupt wider pathophysiological mechanisms[17]. The rTMS, MCS and DBS studies[11,26,41,52,56,58,67,71,84,99] included in this review primarily recruited participants who had medication refractory CPSP which illustrates the effectiveness of these interventions in reducing pain scores. Furthermore, a number of rTMS studies have shown effective neuropathic pain alleviation but were not included in this review as they did not report CPSP-specific data[6,68,69,95,100]. Anaesthetics included in this review also had good effectiveness and, while no side effects were reported, thiamylal[36], morphine[54] and ketamine[80] are known for possible serious adverse side effects, and, therefore, should only be considered for excruciating uncontrollable pain. Thiamylal and ketamine response does appear to predict MCS response, which could be used as a potential test prior to operation, but further research is needed to understand this interaction. Overall, MCS and DBS were found to have similar effectiveness and low number of side effects, but they are invasive treatments and CPSP patients should not consider this as a primary option for treatment. Virtual reality is a treatment that was only explored in one study, but it provided promising pain and physical improvements that should be explored in further studies. It could be a potential add-on therapy where equipment is available. Treatments that require further exploration with more robust methodology include SCS, acupuncture and PNB. Research into potential prophylactic treatments is also needed, as amitriptyline was not found to be effective.

### 4.2 Other Limitations

The overall risk of bias score was “some concerns’’ and GRADE certainty of evidence showed that all included studies had a mean “low” score. Not regarding randomised controlled trials, there are issues in providing full reports on all relevant statistical metrics, disclosing full statistical analysis methodology, reporting side effects, and reporting withdrawal symptoms. VAS assessment scoring was also often not fully disclosed with just a mention that VAS score was taken, and some studies opted for different pain measurements which reduced comparison reliability. Session duration, session length, medication strength, follow-up length, and overall study length also increased heterogeneity and limited generalisability for group meta-analyses. While studies were grouped into either pharmacological, physical, or neuromodulation types for meta-analysis, there was considerable heterogeneity between some intervention modalities. Subgroup analyses provided supplementary validity to whole-group analyses.

There was also high variability in CPSP certainty across studies. Many studies did not perform additional assessments of participants, or relied solely on neuropathic pain questionnaires. However, good quality studies included in this review performed multiple assessments of stroke, exclusion of other conditions, and assessment of pain qualities. Some neuromodulation studies which were performed on primarily thalamic stroke patients, may need to be reassessed with a sample of wider stroke aetiologies. This forms part of a wider group data limitation and strategies to individualise treatments may result in improvements in outcomes.

### 4.3 Conclusions

While there have been other literature reviews and meta-analyses of CPSP treatments[37,94], they either had a focus on just one intervention modality, were smaller in literature search scope, or included only RCT studies[64,37,94]. This review is the most comprehensive overview of different treatment modalities available for CPSP with 16 different treatments in 42 original studies investigating pharmacological, physical and neuromodulation interventions. Using three different analysis approaches – random-effects, proportional and narrative – allowed this review to collate and compare information from a large selection of intervention types and their effectiveness on CPSP pain reduction, secondary outcomes, and associated side effects. While overall quality of studies was moderate to low, there were a number of high quality controlled trials that provide a limited understanding of how different treatments compare. Future research should focus on conducting large sample randomised controlled trials to better understand effectiveness and potential side effects. There is also a lack of longitudinal studies, whether it is randomised controlled trials or retrospective reviews, to understand whether pain control is maintained in the long term and whether it improves over a longer period of time. It is hoped that findings of this review will provide guidance towards future CPSP treatment research and development of a more standardised treatment approach.

## Data Availability

All data produced in the present study are available upon reasonable request to the authors

## Acknowledgments

The authors extend a thank you to the Medical Research Council and Discovery Medicine North (DiMeN) doctoral training programme for funding this research.

There are no conflicts of interest.

## PRISMA 2020 Checklist

| Section and Topic | Item # | Checklist item | Location where item is reported |
| --- | --- | --- | --- |
| **TITLE** |  |  |  |
| Title | 1 | Identify the report as a systematic review. | Title Page |
| **ABSTRACT** |  |  |  |
| Abstract | 2 | See the PRISMA 2020 for Abstracts checklist. | Abstract Page |
| **INTRODUCTION** |  |  |  |
| Rationale | 3 | Describe the rationale for the review in the context of existing knowledge. | Manuscript Pages 1-2 |
| Objectives | 4 | Provide an explicit statement of the objective(s) or question(s) the review addresses. | Manuscript page 2 |
| **METHODS** |  |  |  |
| Eligibility criteria | 5 | Specify the inclusion and exclusion criteria for the review and how studies were grouped for the syntheses. | Manuscript page 5 |
| Information sources | 6 | Specify all databases, registers, websites, organisations, reference lists and other sources searched or consulted to identify studies. Specify the date when each source was last searched or consulted. | Table 1 |
| Search strategy | 7 | Present the full search strategies for all databases, registers and websites, including any filters and limits used. | Manuscript page 5 |
| Selection process | 8 | Specify the methods used to decide whether a study met the inclusion criteria of the review, including how many reviewers screened each record and each report retrieved, whether they worked independently, and if applicable, details of automation tools used in the process. | Manuscript page 5 |
| Data collection process | 9 | Specify the methods used to collect data from reports, including how many reviewers collected data from each report, whether they worked independently, any processes for obtaining or confirming data from study investigators, and if applicable, details of automation tools used in the process. | Manuscript page 5-7 |
| Data items | 10a | List and define all outcomes for which data were sought. Specify whether all results that were compatible with each outcome domain in each study were sought (e.g. for all measures, time points, analyses), and if not, the methods used to decide which results to collect. | Manuscript page 9 |
|  | 10b | List and define all other variables for which data were sought (e.g. participant and intervention characteristics, funding sources). Describe any assumptions made about any missing or unclear information. | Manuscript page 9 |
| Study risk of bias assessment | 11 | Specify the methods used to assess risk of bias in the included studies, including details of the tool(s) used, how many reviewers assessed each study and whether they worked independently, and if applicable, details of automation tools used in the process. | Manuscript page 7 |
| Effect measures | 12 | Specify for each outcome the effect measure(s) (e.g. risk ratio, mean difference) used in the synthesis or presentation of results. | Manuscript page 9 |
| Synthesis methods | 13a | Describe the processes used to decide which studies were eligible for each synthesis (e.g. tabulating the study intervention characteristics and comparing against the planned groups for each synthesis (item #5)). | Manuscript page 9 |
|  | 13b | Describe any methods required to prepare the data for presentation or synthesis, such as handling of missing summary statistics, or data conversions. | Manuscript page 9 |
|  | 13c | Describe any methods used to tabulate or visually display results of individual studies and syntheses. | Manuscript page 9 |
|  | 13d | Describe any methods used to synthesize results and provide a rationale for the choice(s). If meta-analysis was performed, describe the model(s), method(s) to identify the presence and extent of statistical heterogeneity, and software package(s) used. | Manuscript page 9 |
|  | 13e | Describe any methods used to explore possible causes of heterogeneity among study results (e.g. subgroup analysis, meta-regression). | Manuscript page 9 |
|  | 13f | Describe any sensitivity analyses conducted to assess robustness of the synthesized results. | Manuscript page 9 |
| Reporting bias assessment | 14 | Describe any methods used to assess risk of bias due to missing results in a synthesis (arising from reporting biases). | Manuscript page 9 |
| Certainty | 15 | Describe any methods used to assess certainty (or confidence) in the body of evidence for an outcome. | Manuscript page 7- |

PRISMA 2020 Checklist
| Section and Topic | Item # | Checklist item | Location where item is reported |
| --- | --- | --- | --- |
| assessment |  |  | 9 |
| **RESULTS** |  |  |  |
| Study selection | 16a | Describe the results of the search and selection process, from the number of records identified in the search to the number of studies included in the review, ideally using a flow diagram. | Page 9, Figure 1 |
|  | 16b | Cite studies that might appear to meet the inclusion criteria, but which were excluded, and explain why they were excluded. | Page 9, Figure 1 |
| Study characteristics | 17 | Cite each included study and present its characteristics. | Table 4, Supplementary table 1 |
| Risk of bias in studies | 18 | Present assessments of risk of bias for each included study. | Table 5 |
| Results of individual studies | 19 | For all outcomes, present, for each study: (a) summary statistics for each group (where appropriate) and (b) an effect estimate and its precision (e.g. confidence/credible interval), ideally using structured tables or plots. | Pages 17-22, Figure 2-8 |
| Results of syntheses | 20a | For each synthesis, briefly summarise the characteristics and risk of bias among contributing studies. | Pages 17-22, Figure 2-8 |
|  | 20b | Present results of all statistical syntheses conducted. If meta-analysis was done, present for each the summary estimate and its precision (e.g. confidence/credible interval) and measures of statistical heterogeneity. If comparing groups, describe the direction of the effect. | Pages 17-22, Figure 2-8 |
|  | 20c | Present results of all investigations of possible causes of heterogeneity among study results. | Pages 17-22, Figure 2-8 |
|  | 20d | Present results of all sensitivity analyses conducted to assess the robustness of the synthesized results. | Pages 17-22, Figure 2-8 |
| Reporting biases | 21 | Present assessments of risk of bias due to missing results (arising from reporting biases) for each synthesis assessed. | Table 5 |
| Certainty of evidence | 22 | Present assessments of certainty (or confidence) in the body of evidence for each outcome assessed. | Table 5 |
| **DISCUSSION** |  |  |  |
| Discussion | 23a | Provide a general interpretation of the results in the context of other evidence. | Page 23-26 |
|  | 23b | Discuss any limitations of the evidence included in the review. | Page 25 |
|  | 23c | Discuss any limitations of the review processes used. | Page 25 |
|  | 23d | Discuss implications of the results for practice, policy, and future research. | Page 26 |
| **OTHER INFORMATION** |  |  |  |
| Registration and protocol | 24a | Provide registration information for the review, including register name and registration number, or state that the review was not registered. | CRD42022371835 |
|  | 24b | Indicate where the review protocol can be accessed, or state that a protocol was not prepared. | CRD42022371835 |
|  | 24c | Describe and explain any amendments to information provided at registration or in the protocol. | Amended to include additional databases |
| Support | 25 | Describe sources of financial or non-financial support for the review, and the role of the funders or sponsors in the review. | Funded by Medical Research Council as part of PhD. Stated in |

PRISMA 2020 Checklist
| Section and Topic | Item # | Checklist item | Location where item is reported |
| --- | --- | --- | --- |
|  |  |  | Acknowledgements |
| Competing interests | 26 | Declare any competing interests of review authors. | Stated in Acknowledgements |
| Availability of data, code and other materials | 27 | Report which of the following are publicly available and where they can be found: template data collection forms; data extracted from included studies; data used for all analyses; analytic code; any other materials used in the review. | none |

From: Page MJ, McKenzie JE, Bossuyt PM, Boutron I, Hoffmann TC, Mulrow CD, et al. The PRISMA 2020 statement: an updated guideline for reporting systematic reviews. BMJ 2021;372:n71. doi: 10.1136/bmj.n71
For more information, visit: http://www.prisma-statement.org/

## Notes

### Competing Interest Statement

The authors have declared no competing interest.

### Clinical Protocols

https://www.crd.york.ac.uk/prospero/display_record.php?RecordID=371835

### Funding Statement

This study was funded by Medical Research Council as part of Discovery Medicine North PhD Stundetship

