## Supplemental Table 1 for "Management of Central Post-Stroke Pain: Systematic Review and Meta-Analysis"

*Supplementary Table 1 Study Characteristics; Random-Effects meta-analysis in grey, proportional meta-analysis in light blue, and narrative review studies in white*

| **Study ID** | **Methodology**  **T: Type**  **C: Control**  **B: Blinding** | **Intervention**  **T: Type**  **S: Specific Treatment**  **C: Control/Sham type** | **Treatment Specificity**  **S: Strength**  **A: Administration**  **D: Duration** | **N**  **T: Total**  **C: CPSP patients** | **Demographics**  **A: CPSP patient age in years mean (SD/range)**  **G: Gender, N; Male (Female)**  **C: CPSP duration months (SD/range)** | **Reported outcome measures**  **A: Pain**  **B: Psychological**  **C: Physical** |
| --- | --- | --- | --- | --- | --- | --- |
| Bae, 2014[6] | T: Randomised trial  C: Sham trial  B: Single | T: Neuromodulation  S: tDCS^A^  C: Current only applied for 30 seconds | S: 2mA  A: M1^B^  D: 3 times per week for 3 weeks | T: 14  C: 14 | A: 51.70 (RI*)  G: 7 (7)  D: 14.6 (RI*) | A: VAS^1^  B: NR**  C: Skin temperature, QST^2^ |
| Bainton, 1992[7] | T: Randomised trial  C: Crossover  B: Double | T: Pharmacological  S: Naloxone  C: Saline | S: 8mg  A: Injection  D: 45 min to 190 min | T: 20  C: 20 | A: 61.10 (45 – 74)  G: 7 (13)  D: NR** | A: BPI^3^  B: NR**  C: NR** |
| Cho, 2013[15] | T: Randomised trial  C: Placebo-control  B: Single | T: Physical  S: Bee venom acupuncture  C: Saline injection in to 1 acupoint | S: 0.05ml diluted bee venom  A: acupoints of affected side  D: Once | T: 16  C: 16 | A : NR (36 - 88)  G: NR**  D: NR** | A: VAS  B: NR**  C: NR** |
| De Oliveira, 2014[19] | T: Randomised trial  C: Sham-control  B: Double | T: Neuromodulation  S: rTMS^C^  C: Identical sham coil emitting sound | S: 1250 pulses at 10 Hz, 50% MT^D^  A: Premotor cortex  D: RI* | T: 21  C: 21 | A: 56.33 ( 10.59)  G: 11 (10)  D: 57.40 (50.24) | A: VAS, NPS^4^  B: HAM-A^5^, HAM-D^6^  C: NR** |
| Kim, 2011[42] | T: Randomised trial  C: Placebo control  B: Double | T: Pharmacological  S: Pregabalin  C: Placebo pill | S: Mean 356.8 mg (125 – 539.70)  A: Daily pill  D: 4 weeks | T: 219  C: 219 | A: 58.28 (RI)  G: 137 (82)  D: 28.2 (1.20 – 212.40) | A: VAS^1^  B: HADS^7^  C: NR** |
| Mahesh, 2022[57] | T: Randomised trial  C: Placebo control  B: Double | T: Pharmacological  S: Duloxetine  C: Placebo pill | S: 30mg to 60mg (based on NRS^8^ pain intensity at 2 weeks)  A: Daily pill  D: 4 weeks | T: 82  C: 82 | A: 55.80 (9.90)  G: 47 (35)  D: 2 (6.67 - 48) | A: NRS^8^, SF-MPQ^9^, PDI^10^  B: PGIC^11^  C: NR** |
| McGeoch, 2008[59] | T: Cross sectional Within-subject  C: Sham-control  B: Single | T: Neurostimulation  S: Vestibular stimulation  C: Body temperature irrigation or ice pack application to the pinna | S: Cold water  A: Injection in to air canal  D: 1 session | T: 9  C: 9 | A: 60.22 (15.72)  G: 3 (6)  D: NR (30-180) | A: NRS^8^  B: NR**  C: NR** |
| O’Neill, 2018[65] | T: Randomised trial  C: Crossover  B: Double | T: Neuromodulation  S: tDCS^A^  C: Current only applied for 5 seconds | S: 1.4mA  A: M1^B^  D: Daily for 5 days | T: 24  C: 9 | A: NR**  G: 7 (2)  D: NR** | A: NRS^8^  B: HADS^7^  C: NR** |
| Ojala, 2021[68] | T: Randomised trial  C: Crossover  B: Double | T: Neuromodulation  S: rTMS^C^  C: non-conductive plastic cover | S: 5050 pulses at 10Hz, 90% of MT^D^  A: M1^B^ and S2^E^ as separate sessions  D: 2 weeks per crossover session | T: 17  C: 17 | A: 58.80 (7.10)  G: 8 (9)  D: 67.2 (38.4) | A: NRS^8^  B: BDI^12^, PASS-20^13^, EQ-5D-3L^14^  C: DASH^15^ |
| Onouchi, 2014[69] | T: Open-label  C: Comparison  B: None | T: Pharmacological  S: Pregabalin  C: Another pain group | S: 300mg to 600mg  A: Daily pill  D: 52 weeks | T: 100  C: 60 | A: 61.70 (8.50)  G: 42 (18)  D: NR** | A: BPI^16^, VAS^1^, SF-MPQ^9^  B: NR**  C: NR** |
| Xiao-nong, 2012[95] | T: Crossover  C: Comparison trial  B: None | T: Physical  S: Acupuncture  C: Western Medicine | S: Puncture of acupoints 8-12 mm to 20-40mm in depth, 0.4g to 1.2g of carbamazepine  A: NR**  D: 15 minutes | T: 11  C: 11 | A: NR**  G: 4 (2)  D: NR** | A: VAS^1^  B: NR**  C: NR** |
| Zhao, 2021[100] | T: Randomised trial  C: Sham-control  B: Double | T: Neuromodulation  S: rTMS^C^  C: Identical coil with sound but no stimulation | S: 1500 pulses at 10HZ, 80% of MT^D^  A: M1^B^  D: 6 days per week for 3 weeks | T: 38  C: 38 | A: 49.55 (11.29)  G: 19 (19)  D: 6.24 (RI*) | A: NRS^8^, SF-MPQ-CN^17^  B: HAM-A^5^, HAM-D^6^  C: NR** |
| Boccard, 2012[11] | T: Cross sectional  C: None  B: None | T: Neuromodulation  S: DBS^F^  C: None | S: 5 to 50Hz, pulse width 200 to 450ms, amplitude 0.5 to 5V  A: PVG^G^, VPL^H^, VPM^I^  D: 3 months | T: 85  C: 23 | A: 58.8 (9.10)  G: 14 (2)  D: NR** | A: VAS^1^, MPQ^18^  B: EQ-5D^14^  C: SF-36^19^ |
| Choi, 2021[16] | T: Retrospective  C: None  B: None | T: Neuromodulation  S: PNB^J^  C: None | S: 3 ampoules of 1.8 ml solution each with 36 mg/1.8 ml lidocaine HCl and 0.009 mg/1.8 ml epinephrine  A: Ulnar nerve blocks in proximal forearm or femoral, obturator and sciatic nerve blocks in proximal thigh  D: 1 day | T: 22  C: 22 | A: 56.27 (13.33)  G: 8 (14)  D: 11.40 (16.66) | A: BPI^16^  B: NR**  C: NR** |
| Gou, 2022[26] | T: Retrospective  C: None  B: None | T: Neuromodulation  S: MCS^K^  C: None | S: 30-50 HZ, pulse width 210–300 μs at 3.5 – 7.0 V  A: M1^B^  D: 5-7 days | T: 21  C: 21 | A: 58.52 (7.27)  G: NR**  D: 34.38 (28.39) | A: VAS^1^, NPSI^20^  B: PSQI^21^  C: NR** |
| Kim, 2019[43] | T: Cross sectional  C: None  B: None | T: Pharmacological  S: Duloxetine  C: None | S: 30mg-60mg once daily  A: Daily pill  D: 3 weeks | T: 37  C: 37 | A: 48.90 (12.10)  G: 16 (21)  D: 37.2 (49.2) | A: NRS^8^, SF-MPQ^17^  B: NR**  C: NR** |
| Lefaucheur, 2004[52] | T: Randomised within-subjects trial  C: Sham-control  B: Single | T: Neuromodulation  S: rTMS^C^  C: Magstim placebo coil | S: 20 pulses of 5 seconds at 10HZ, 80% MT^D^  A: M1^B^  D: NR | T: 60  C: 12 | A: RI*  G: RI*  D: NR** | A: VAS^1^  B: NR**  C: NR** |
| Lin, 2018[56] | T: Cross sectional  C: None  B: None | T: Neuromodulation  S: rTMS^C^  C: None | S: 1000 pulses at 10HZ daily, 90% MT^D^  A: M1^B^  D: 10 days | T: 7  C: 7 | A: 53.57 (7.16)  G: 5 (2)  D: 44.4 (6-132) | A: VAS^1^  B: HAM-A^5^, HAM-D^6^  C: Laser Evoked Potentials |
| Matsumura, 2012[58] | T: Within-subjects  C: Placebo control  B: NR | T: Neuromodulation  S: rTMS^C^  C: Coils elevated at an angle of 45° from the skull | S: 500 pulses at 5 Hz, 100% MT^D^  A: M1^B^  D: 1 day | T: 20  C: 20 | A: 63.60 (8.10)  G: 12 (8)  D: 38.15 (6 –190) | A: VAS^1^, PGIC^11^  B: NR**  C: NR** |
| Mohamed, 2010[62] | T: Retrospective  C: None  B: None | T: Neuromodulation  S: SCS^L^  C: None | S: RI*  A: Spinal level C4 to C7 for upper limb pain or T9 to T12 for lower limb  D: 2-7 days trial, then permanent | T: 30  C: 30 | A: 64.80 (7.40)  G: 21 (9)  D: 44.80 (6 – 156) | A: VAS^1^  B: NR**  C: NR** |
| Ohn, 2012[67] | T: Cross sectional  C: None  B: None | T: Neuromodulation  S: rTMS^C^  C: None | S: 1000 pulses at 10HZ, 90% MT^D^  A: M1^B^  D: 5 times per day for 5 days | T: 22  C: 22 | A: 54.90 (9.00)  G: 13 (9)  D: 21.90 (17.20) | A: VAS^1^  B: HAM-D^6^  C: NR** |
| Owen, 2006[71] | T: Cross sectional  C: None  B: None | T: Neuromodulation  S: DBS^F^  C: None | S: NR  A: VPL^H^, PVG^G^  D: 1 week | T: 15  C: 15 | A: 58.60 (37 - 74)  G: 12 (3)  D: 62.4 (NR) | A: VAS^1^, MPQ^17^  B: NR**  C: NR** |
| Quesada, 2019[74] | T: Randomised trial  C: Sham-control  B: Double | T: Neuromodulation  S: rTMS^C^  C: Sham train side of coil | S: 1600 pulses at 20 Hz, 80% MT^D^  A: M1^B^  Dtab;e: 27 minute session | T: 42  C: 19 | A: RI*  G: RI*  D: RI* | A: VAS^1^, NPSI  B: EQ-5D^20^  C: NR** |
| Shimodozono, 2009[79] | T: Cross sectional  C: None  B: None | T: Pharmacological  S: Fluvoxamine  C: None | S: 25mg to 125mg  A: Daily pill  D: Daily for 2 to 4 weeks | T: 28  C: 28 | A: 62.20 (9.70)  G: 13 (15)  D: NR** (1-108) | A: VAS^1^  B: SDS^22^  C: NR** |
| Tanei, 2019[84] | T: Retrospective  C: None  B: None | T: Neuromodulation  S: SCS^L^  C: None | S: 30Hz with pulse width 240 μs  A: Midline ipsilateral to area of pain  D: 12 months | T: 18  C: 18 | A: 63.90 (8.80)  G: 10 (8)  D: 54 (43.20) | A: VAS^1^  B: NR**  C: NR** |
| Zhang, 2018[99] | T: Cross sectional  C: None  B: None | T: Neuromodulation  S: MCS^K^  C: None | S: 30-50 HZ, pulse width 210–300 μs at 3.5 – 7.0 V  A: M1^B^  D: 5-7 days | T: 16  C: 16 | A: 59.9 (7.80)  G: 8 (8)  D: NR** | A: VAS^1^, NPSI^20^  B: NR**  C: NR** |
| Hasan, 2013[27] | T: Cross sectional  C: None  B: None | T: Neuromodulation  S: rTMS^C^  C: None | S: 2000 pulses at 10 Hz, 80-90% MT ^D^  A: M1 ^B^  D: 23 minute sessions ever 3 – 5 days for 21 day cycle | T: 14  C: 14 | A: RI*  G: 10 (4)  D: NR** | A: BPI^16^  B: PGIC^11^, HADS^7^  C: NR** |
| Jungehulsing, 2013[38] | T: Randomized cross over  C: Placebo control  B: Double | T: Pharmacological  S: Levetiracetam  C: Placebo pill | S: 2130 mg (SD 830)  A: Daily pill  D: 8 weeks per cross over phase | T: 42  C: 42 | A: 61.50 (50 – 76)  G: 26 (16)  D: 48 (4.8 – 132) | A: MPQ^9^  B: BDI^12^  C: QST^23^ |
| Kalita, 2017[39] | T: Cross over  C: Comparison trial  B: None | T: Pharmacological,  S: Pregabalin, Lamotrigine  C: None | S: Lamortrigine 200mg, Pregabalin 600mg  A: Daily pill  D: 12 weeks per trial | T: 30  C: 30 | A: 54.50 (25 – 74)  G: 26 (4)  D: 8.35 (16.19) | A: VAS^1^  B: HADS^7^  C: QST^23^ |
| Katayama, 1998[40] | T: Cross sectional  C: None  B: None | T: Neuromodulation  S: MCS^K^  C: None | S: 25 – 50 Hz at 2 – V  A: M1 ^B^  D: 1 week | T: 31  C: 31 | A: RI*  G: 20 (11)  D: NR** | A: RI*  B: NR**  C: NR** |
| Khedr, 2005[41] | T: Between-subjects  C: Placebo control  B: Single | T: Neuromodulation  S: rTMS^C^  C: Coil elevated and angled away from the head | S: 2000 pulses at 20HZ, 80% MT ^D^  A: M1 ^B^  D: 10 minutes per day for 5 days | T: 58  C: 24 | A: 52.30 (10.30)  G: NR**  D: 18 (17) | A: VAS^1^, LANSS^24^  B: NR**  C: NR** |
| Kobayashi, 2014[46] | T: Cross sectional  C: Sham-control  B: Single | T: Neuromodulation  S: rTMS^C^  C: Coil elevated and angled away from the head | S: 500 pulses at 5 Hz, 90% MT ^D^  A: M1 ^B^  D: 10 minutes | T: 18  C: 18 | A: 64 (9.90)  G: 12 (6)  D: 9 (2 – 26) | A: VAS^1^  B: NR**  C: NR** |
| Lampl, 2002[50] | T: Randomized trial  C: Placebo control  B: Double | T: Pharmacological  S: Amitriptyline  C: Placebo pill | S: 75mg  A: Daily pill  D: 3 weeks | T: 39  C: 39 | A: 60.50 (RI*******)  G: RI*  D: RI* | A: NR**  B: NR**  C: NR** |
| Leijon, 1989[53] | T: Randomized cross over  C: Comparison trial  B: None | T: Pharmacological  S: Amitriptyline, Carbamazepine  C: None | S: Amitriptyline – 75mg, Carbamazepine – 800mg  A: Daily pill  D: 4 weeks | T: 15  C: 15 | A: 66 (53 - 74)  G: 12 (3)  D: 54 (11 - 154) | A: 10-step verbal scale  B: CPRS^25^  C: NR** |
| McLean, 2018[60] | T: Cross sectional  C: None  B: None | T: Neuromodulation  S: rTMS^C^  C: None | S: 10 trains at 10 Hz, 80% MT ^D^  A: M1 ^B^  D: daily for 11 days | T: 50  C: 5 | A: RI*  G: RI*  D: RI* | A: VAS^1^  B: NR**  C: NR** |
| Mohamed, 2010[62] | T: Retrospective  C: None  B: None | T: Neuromodulation  S: SCS^L^  C: None | S: RI*  A: Spinal level C4 to C7 for upper limb pain or T9 to T12 for lower limb  D: 2-7 days trial, then permanent | T: 30  C: 30 | A: 64.80 (7.40)  G: 21 (9)  D: 44.80 (6 – 156) | A: VAS^1^  B: NR**  C: NR** |
| Petramfar, 2010[73] | T: Retrospective  C: None  B: none | T: Pharmacological  S: Lamotrigine  C: None | S: 25mg rising to 100mg over 5 weeks and then maintained  A: Daily pill  D: 24 weeks | T: 17  C: 17 | A: 60.20 (12.40)  G: 7 (10)  D: 8.40 (4 – 20) | A: BPI^16^  B: NR**  C: NR** |
| Rahajeng, 2018[76] | T: Observational  C: None  B: None | T: Pharmacological  S: Pregabalin  C: None | S: 75 mg  A: Daily pill  D: 12 weeks | T: 36  C: 36 | A: 57.11 (7.51)  G: 21 (15)  D: NR** | A: BPI^16^ short form  B: NR**  C: NR** |
| Simmonds, 2016[82] | T: Within-subjects  C: Placebo control  B: None | T: Physiological  S: VR^D^  C: No VE^M^ and Neutral VE^M^ conditions | S: “Snow World” and “Hot World” virtual environments  A: Head mounted display  D: 3-5 minutes per condition | T: 12  C: 12 | A: 61 (7.02)  G: 7 (5)  D: NR** | A: VAS^1^  B: Mood  C: QST^23^ |
| Tsubokawa, 1993[86] | T: Cross sectional  C: None  B: None | T: Neuromodulation  S: Cortex stimulation  C: None | S: 5 to 120 HZ  A: VPL^H^, IC^N^, ML^O^, PAG^P^, PBR^R^  D: Implanted indefinitely | T: 11  C: 11 | A: 58.90 (52 – 72)  G: 4 (7)  D: NR** (12-48) | A: VAS^1^  B: NR**  C: NR** |
| Vestergaard, 2001[91] | T: Randomized trial  C: Cross over  B: Double | T: Pharmacological  S: Lamotrigine  C: Placebo pill | S: 200 mg  A: Daily pill  D: 8 weeks per treatment period | T: 30  C: 30 | A: 59 (37 – 77)  G: 18 (12)  D: 24 (3.6 – 144) | A: VAS^1^, evoked pain  B: NR**  C: QST^23^ |
| Yamamoto, 1997[97] | T: Cross sectional  C: Comparison trial  B: Single (pharmacological) None (MCS^K^) | T: Neuromodulation, Pharmacological  S: MCS^K^, morphine, thiamylal, keramine  C: Saline (pharmacological) | S: MCS 25 – 50 Hz at 2-5 V; morphine 18 mg, thiamylal 250mg, ketamine 25mg  A: M1^B^ (MCS^K^); injection (pharmacological)  D: MCS^K^ 15 – 30 min; morphine 30 min, thiamylal 25 min, ketamine 25 min | T: 39  C: 39 | A: 58.70 (35 – 72)  G: 25 (14)  D: 8.90 (1 – 6) | A: VAS^1^  B: NR**  C: NR** |

**A** Transcranial Direct Current Stimulation; **B** Primary Motor Cortex; **C** Repetitive Transcranial Magnetic Stimulation; **D** Motor threshold; **E** Secondary somatosensory cortex; **F** Deep Brain Stimulation; **G** Periventricular grey; **H** Ventral posterolateral nucleus (thalamus); **I** Ventral Posteromedial nucleus; **J** Peripheral Nerve Block; **K** Motor Cortex Stimulation; **L** Spinal Cord Stimulation; **M** Virtual Environment; **N** Internal Capsule; **O** Medial Lemniscus; **P** Periaqueductal Gray; **R** Pontomesencephalic Parabrachial Region;

**1** Visual Analogue Scale; **2** Quantitative Sensory Testing; **3** Brief Pain Inventory (0 to 10); **4** Neuropathic Pain scale; **5** Hamilton Rating Scale – Anxiety; **6** Hamilton Rating Scale – Depression; **7** Hospital Anxiety and Depression Scale; **8** Numeric Rating Scale (0 to 100); **9** Short-form McGill Pain Questionnaire; **10** Pain Disability Index; **11** Patient Global Impression of Change; **12** Beck Depression Inventory; **13** Pain Anxiety Symptoms Scale; **14** European Quality of Life 5 Dimensions 3 Level Version; **15** Disabilities of the Arm, Shoulder, and Hand; **16** Brief Pain Inventory (0 to 10); **17** SF-MPQ Mandarin Chinese version; **18** McGill Pain Questionnaire; **19** Short Form 36-Item Survey Instrument; **20** Neuropathic Pain Symptom Inventory; **21** Pittsburgh Sleep Quality Index; **22** Self-rating Depression Scale; **23** Quantitative Sensory Testing **24** Leeds Assessment Of Neuropathic Symptoms And Signs; **25** Comprehensive Psychopathological Rating Scale; *Reporting Indiscernible – not possible to extract exact numerical value; **Not Reported;
